# Immunological and Antigenic Signatures Associated with Chronic Illnesses after COVID-19 Vaccination

**DOI:** 10.1101/2025.02.18.25322379

**Authors:** Bornali Bhattacharjee, Peiwen Lu, Valter Silva Monteiro, Alexandra Tabachnikova, Kexin Wang, William B. Hooper, Victoria Bastos, Kerrie Greene, Mitsuaki Sawano, Christian Guirgis, Tiffany J. Tzeng, Frederick Warner, Pavlina Baevova, Kathy Kamath, Jack Reifert, Danice Hertz, Brianne Dressen, Laura Tabacof, Jamie Wood, Lily Cooke, Mackenzie Doerstling, Shadan Nolasco, Amer Ahmed, Amy Proal, David Putrino, Leying Guan, Harlan M. Krumholz, Akiko Iwasaki

**Author notes:** Indicates equal contribution (Bornali Bhattacharjee, Peiwen Lu, Valter Silva Monteiro, Alexandra Tabachnikova, Kexin Wang, William B. Hooper, Victoria Bastos, Kerrie Greene).

## Abstract

COVID-19 vaccines have prevented millions of COVID-19 deaths. Yet, a small fraction of the population reports a chronic debilitating condition after COVID-19 vaccination, often referred to as post- vaccination syndrome (PVS). To explore potential pathobiological features associated with PVS, we conducted a decentralized, cross-sectional study involving 42 PVS participants and 22 healthy controls enrolled in the Yale LISTEN study. Compared with controls, PVS participants exhibited differences in immune profiles, including reduced circulating memory and effector CD4 T cells (type 1 and type 2) and an increase in TNFα+ CD8 T cells. PVS participants also had lower anti-spike antibody titers, primarily due to fewer vaccine doses. Serological evidence of recent Epstein-Barr virus (EBV) reactivation was observed more frequently in PVS participants. Further, individuals with PVS exhibited elevated levels of circulating spike protein compared to healthy controls. These findings reveal potential immune differences in individuals with PVS that merit further investigation to better understand this condition and inform future research into diagnostic and therapeutic approaches.

## INTRODUCTION

The rapid development and deployment of COVID-19 vaccines have been pivotal in mitigating the impact of the pandemic^1^. These vaccines have significantly reduced severe illness and mortality associated with SARS-CoV-2 infection^2^. Additionally, vaccinated individuals experience a lower incidence of post-acute sequelae of COVID-19 (PASC) or long COVID, thus highlighting an additional potential benefit of receiving the COVID-19 vaccines ^3, 4^. However, COVID-19 vaccines are associated with rare acute adverse events ^5^ such as myocarditis and pericarditis ^6^, thrombosis and thrombocytopenia^7^, Guillain–Barre syndrome, transverse myelitis, and Bell’s Palsy^8,9^.

In addition, some individuals have reported post-vaccination symptoms resembling long COVID beginning shortly after vaccination. This condition, sometimes referred to as post-vaccination syndrome (PVS) or post-acute COVID-19 vaccination syndrome (PACVS) ^10, 11^, is characterized by symptoms such as exercise intolerance, excessive fatigue, numbness, brain fog, neuropathy, insomnia, palpitations, myalgia, tinnitus or humming in ears, headache, burning sensations, and dizziness^10^. Unlike long COVID, PVS is not officially recognized by health authorities, which has significantly limited patient care and support.

The molecular mechanisms of PVS remain largely unknown. However, there is considerable overlap in self-reported symptoms between long COVID and PVS, as well as shared exposure to SARS-CoV-2 spike (S) protein in the context of inflammatory responses during infection or vaccination^10,12,13^. In susceptible individuals, vaccines may contribute to long-term symptoms by multiple mechanisms. For example, vaccine components, such as mRNA, lipid nanoparticles, and adenoviral vectors, trigger activation of pattern recognition receptors^14,15^. Thus, unregulated stimulation of innate immunity could lead to chronic inflammation. Secondly, it has been shown that the S protein expressed following BNT162b2 or mRNA-1273 vaccination circulates in the plasma as early as one day after vaccination^16,17^. Interaction with full-length S, its subunits (S1, S2), and/or peptide fragments with host molecules may result in prolonged symptoms in certain individuals^16^. Recently, a subset of non-classical monocytes has been shown to harbor S protein in patients with PVS^18^. Further, biodistribution studies on mRNA–LNP platforms in animal models indicate its ability to cross the blood-brain barrier, and the local S expression could result in neurocognitive symptoms ^19, 20^. Third, vaccine-induced immune responses may be triggering the stimulation of autoreactive lymphocytes^21^.

To investigate immunological features in people suffering from persistent symptoms after COVID-19 vaccination, a cross-sectional case-control study was undertaken to identify the immunological correlates of PVS. A total of 42 participants with PVS who had no pre-existing comorbidities and 22 contemporaneous healthy controls who did not report PVS after receiving COVID-19 vaccines were included. An important factor to evaluate was the possibility that PVS might result from an undiagnosed, asymptomatic SARS-CoV-2 infection coinciding with the vaccination period, instead of being directly caused by the vaccine administration. In addition, infection with SARS-CoV-2 significantly impacts immune signatures^22^. Our objectives were twofold: (1) to conduct a two-group case-control analysis of the immunophenotypic profiles of individuals with PVS in comparison with asymptomatic vaccine recipients, and (2) to compare the immunophenotypic profiles of those with PVS with or without a history of SARS-CoV-2 infection. To achieve these, we profiled circulating immune cell populations, antibody responses, and circulating immune modulator levels in addition to assessing the demographic and general health characteristics of the participants.

## RESULTS

### Cohort Description

All the blood samples were collected between December 2022 and November 2023 from the Listen to Immune, Symptom and Treatment Experiences Now (LISTEN) study^23^. The PVS cohort consisted of a total of 42 participants, including 29 females and 13 males with no preexisting comorbidities, whereas the control cohort consisted of 22 participants, including 13 females and 9 males (Figures S1A and 1A). Upon recruitment of 44 PVS participants, two had to be excluded from the analyses due to evidence of pharmacological immunosuppression. Information on index vaccine types was reported by 39 out of 42 PVS participants included in the analyses, and they were Comirnaty (Pfizer) (n=14), Spikevax (Moderna) (n=21), and Jcovden (J&J) (n=4). The most frequent symptoms reported by participants were excessive fatigue (85%), tingling and numbness (80%), exercise intolerance (80%), brain fog (77.5%), difficulty concentrating or focusing (72.5%), trouble falling or staying asleep (70%), neuropathy (70%), muscle aches (70%), anxiety (65%), tinnitus (60%) and burning sensations (57.5%). Further, pairwise Euclidean distances were calculated in a sex-segregated manner based on the presence of symptoms at recruitment and two distinct clusters of symptoms were identified in both (Table S1, Figure S1B).

Between the case and control cohorts, a total of 15 (35.7%) and 10 (45.5%) reported having a history of one or more previous SARS-CoV-2 infections, respectively (Table S2). However, upwards of 40% of SARS-CoV-2 infections are asymptomatic^24^. To further investigate prior history of SARS-CoV-2 infections, plasma specimens were analyzed using the EUA-cleared Elecsys® anti-SARS-CoV-2 immunoassays, which measure the presence of high affinity IgM, IgA, and IgG anti-N antibodies. A cut-off index ≥ 1 was defined as reactive based on previous literature^25^. In the non-reactive group, the antibody indices varied between 0.09 and 0.17, whereas in the reactive group, they ranged between 1.37 and 94.4. Among participants with PVS, 26 (61.9%) were found to be reactive compared with 10 (45.5%) among controls. One participant from each cohort with self-reported history of infection had non-reactive test results. Based on both self-reports and serological analyses the two cohorts were further classified into four subgroups, PVS with no history of infection (PVS-I, n= 15), PVS with a history of infection (PVS+I, n= 27), controls with no history of infection (Control-I, n= 11) and controls with a history of infection (Control+I, n= 11) (Figure 1A, Table S2). Even though all PVS participants developed chronic symptoms following vaccination and not infection, it was important to consider the impact of a subsequent SARS-CoV-2 infection on immune phenotypes analyzed in this study.

**Figure 1:**
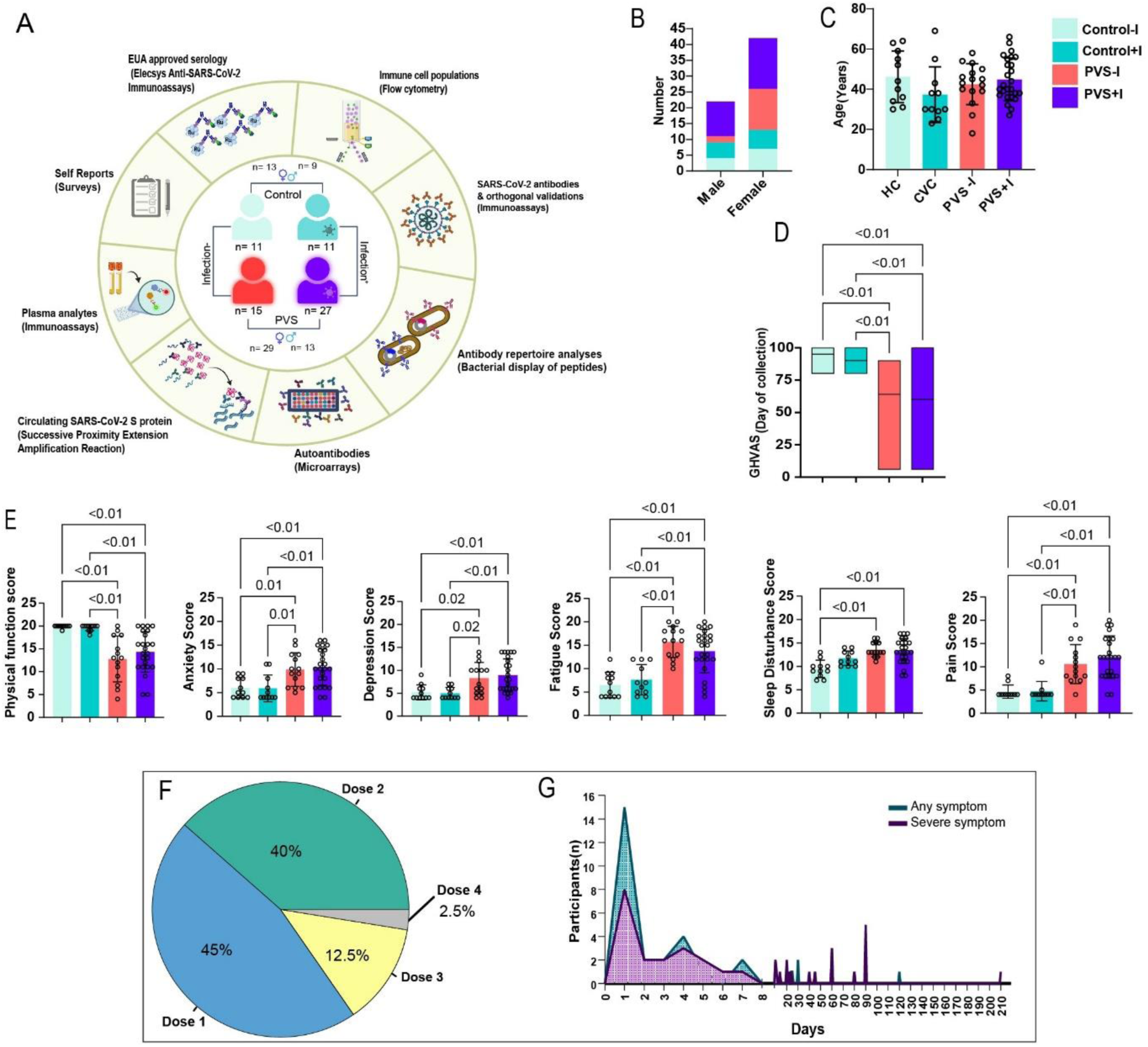
Study overview and cohort information. **A.** Summary of the study design, cohorts and assays included in this study **B.** Stacked bar plots showing the number of males and females in the four subgroups. **C.** Scatter plots showing the distribution of age across subgroups. **D.** Boxplots showing self-reported General Health Visual Analogue Scales (GHVAS) scores self-reported on the day of collection. **E.** Scatter plots showing the Patient-Reported Outcomes Measurement Information System 29-Item Profile Measure (PROMIS-29 v2.0) score differences across domains. Differences in biological sex among the four groups was assessed using Fisher’s exact test. For the rest, significance was assessed using either Mann-Whitney U and Kruskal-Wallis tests with Benjamini–Hochberg false-discovery rate (FDR) correction for multiple comparisons wherever necessary. Only significant differences are highlighted (p values ≤ 0.05). **F.** A pie chart showing the proportion of index vaccine doses within the PVS cohort. **G.** Lines plots showing the distribution of days post-vaccination that the participants develop any or severe symptoms associated with PVS.

Among the demographic variables, there were no significant differences in the number of males and females between cases and controls (Fisher’s Exact test, p= 0.58) or among the four groups (Kruskal-Wallis test, p = 0.25; Figure 1B). Similarly, no significant age differences were observed between cases and controls, median age (42.5 years, PVS; 38 years, controls, Mann-Whitney U test p= 0.27) and among the four groups (p = 0.17; Figure 1C).

The self-reported General Health Visual Analogue Scale (GHVAS) scores on the day of biospecimen collection differed significantly among the four groups (Kruskal-Wallis test, p = <0.01). The controls in both subgroups had significantly higher median scores compared with the PVS subgroups (64, PVS-I; 60, PVS+I; 95, Control-I; 90, Control+I; Figure 1D). The PROMIS-29 physical function, fatigue, pain interference, depression, anxiety, sleep disturbance, and pain interference scores were compared independently among the four groups to gauge the physical and mental health status of the participants. The physical function scores were significantly higher among the controls than the cases irrespective of infection status, median scores (13, PVS-I; 14.5, PVS+I; 20, Control-I; 20, Control+I). The anxiety (9.5, PVS-I; 10, PVS+I; 5, Control-I; 4, Control+I), depression (8.5, PVS-I; 8, PVS+I; 4, Control-I; 4, Control+I), fatigue (16, PVS-I; 15, PVS+I; 6, Control-I; 7, Control+I) and pain scores (9.5, PVS-I; 12, PVS+I;4, Control-I; 4, Control+I) were significantly lower in controls compared with participants with PVS irrespective of infection status. Further, significantly higher sleep disturbance scores were observed only among infection-negative cases compared to control-I participants (13, PVS-I; 10, Control-I) (Figure 1E).

Most individuals in each cohort completed the primary series of vaccines based on WHO recommendations (83.3%, PVS; 100%, Controls; Fisher Exact test, p = 0.09). Participants with PVS received significantly fewer COVID-19 vaccine doses compared with controls, median vaccine numbers (2, PVS; 4, controls; Fisher Exact test, p = <0.01). On similar lines, the median number of days post the latest vaccination was significantly higher among cases with a median of 585 days (±190) compared with 199 days (±217) among controls (Mann-Whitney U test, p= <0.01). In 85% of the cases, participants identified the index vaccine dose as being part of the primary series [dose 1(45%) and dose 2(40%); Figure 1F]. The median number of days for the development of any symptom was 4 [Interquartile range (IQR): 23 days], while for severe symptoms, it was 10 (IQR: 44 days) post-vaccination. A high proportion of participants with PVS developed any symptoms (70%) or severe symptoms (52.2%) within 10 days of vaccination (Figure 1G).

### Differences in circulating immune cell populations

To determine immune signatures of PVS, peripheral blood mononuclear cells (PBMC) were analyzed using flow cytometry. Among the cell populations of myeloid lineage, proportions of non-classical monocytes (CD14^low^CD16^high^; Mann-Whitney U test, p= 0.03) were significantly higher in the PVS cohort compared to the controls without significant differences in the percentage of total monocytes despite greater median values in PVS (Figure 2A). The median percentage of conventional type 2 dendritic cells (cDC2; CD304^-^/HLA-DR^+^/CD1c^+^) was significantly lower among the participants with PVS compared to the controls (p= 0.02) while no differences were observed in the proportions of conventional type 1 dendritic cells (cDC1; (CD304^-^/HLA-DR^+^/CD141^+^) (Figure 2B). Pairwise comparisons were also executed to understand the differences among the PVS subgroups with or without a history of infection. Among the low-density granulocytes, no differences were observed between cases and controls in the proportions of eosinophils (CD66b^+^CD56^-^CD16^-^) or between the cases and controls in the infected or uninfected subgroups but the proportion of neutrophils (CD66b^+^/CD56^-^ /CD16^+^) was significantly higher (p= 0.02) in infection positive PVS subgroup (PVS+I) compared to the convalescent controls (control+I) (Figure S2A). Significantly lower and higher proportions of classical and non-classical monocytes, respectively, were observed among the PVS+I compared to the control+I subgroup (p(cMonocytes)= <0.01; p(ncMonocytes)= 0.03) with no differences between the cases and controls without prior history of SARS-CoV-2 infection (Figure S2B). Next, significantly higher proportions of both cDC1 and cDC2 cells were observed in the control+I subgroup compared to the PVS+I subgroup (p= 0.03 and p= <0.01), respectively (Figure S2C).

**Figure 2:**
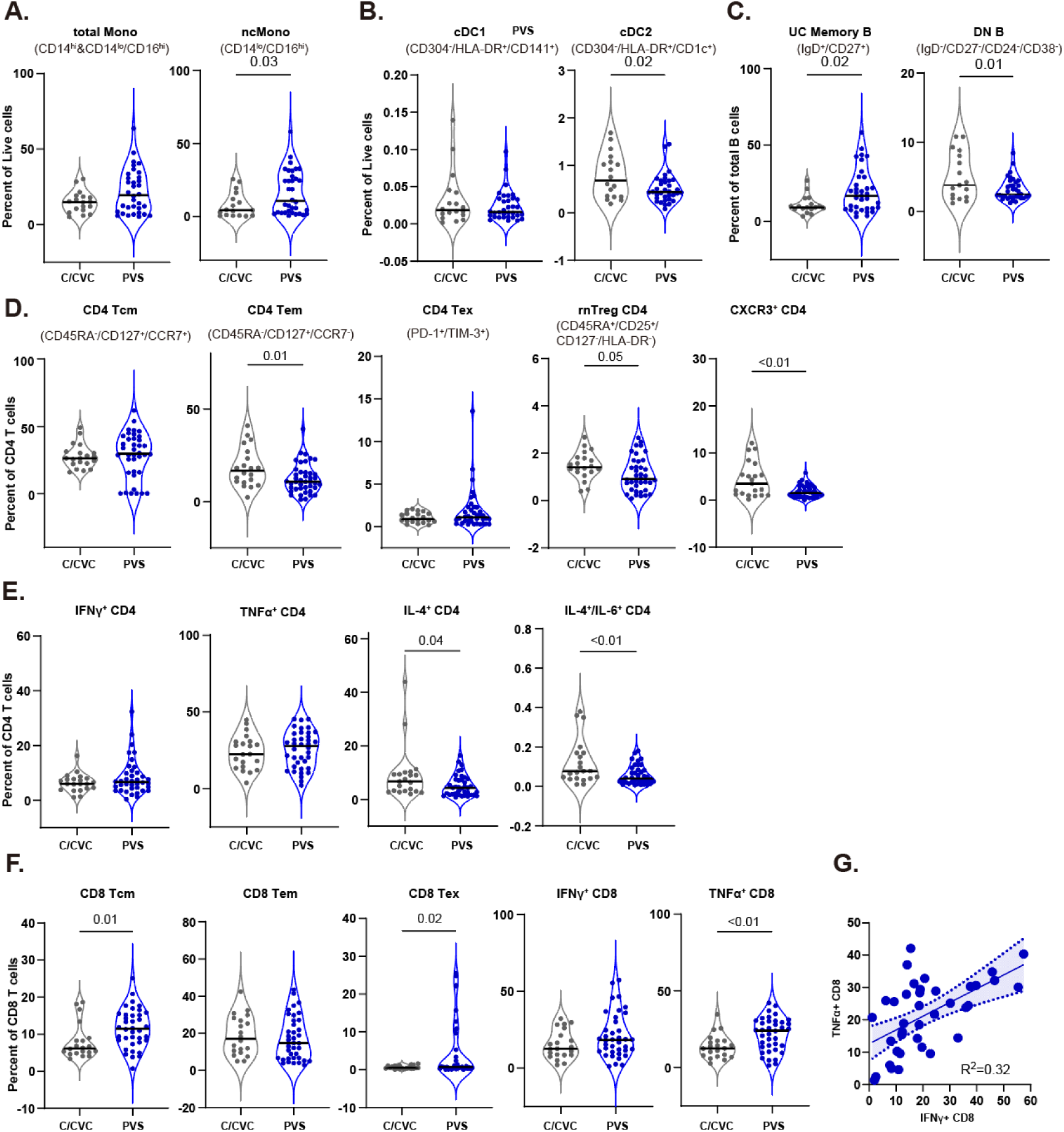
Immune cell feature of myeloid and lymphoid cells in PVS patients. **A-B.** Violin plots of myeloid peripheral blood mononuclear populations (PBMCs) plotted by groups as percentages of respective parent populations (live cells). **C.** Violin plots of B lymphocyte subsets from PBMCs plotted as percentages of respective parent populations (total B cells). **D.** Violin plots of various CD4 T cell subsets. **E.** Violin plots of various cytokine-producing CD4 T cell subsets. **F.** Violin plots of various CD8 T cell subsets and cytokine-producing CD8 T cell subsets. Significance differences were assessed using Mann-Whitney U tests with Benjamini–Hochberg false-discovery rate (FDR) correction for multiple comparisons. **G.** Linear regressions of TNFα producing CD8 T cells and IFNγ producing CD8 T cells. Spearman’s correlation was calculated with corresponding p-values. Dotted lines depict linear regressions, with the area inside representing 95% CI.

Among the B cell populations, relative proportions of unswitched memory B cells (US memory B cells; CD19^+^/CD27^+^/IgD^+^) were significantly higher (p= 0.02) while the proportion of double negative B cells (DN B; IgD^-^/CD27^-^/CD24^-^/CD38^-^) was observed to be lower (p= 0.01) in the PVS cohort compared with controls (Figure 2C).

Significant differences in subsets of circulating immune cell populations were observed across T cell lineages. Upon assessment of the T cell populations, notably higher proportions of effector memory CD4 T cell subsets (CD4^+^Tem; CD45RA^-^/CD127^+^/CCR7^-^; p= 0.01) and resting natural CD4^+^ Treg; CD45RA^+^/CD25^+^/CD127^-^/HLA-DR^-^; p= 0.05) were observed among the controls (Figure 2D). However, the PVS cohort had significantly higher proportions of exhausted CD8 T cell (CD8^+^ Tex; PD-1^+^/TIM3^+^; p= 0.02) (Figure 2F) with no observed differences in the CD4^+^ central memory (CD4^+^ cm; CD45RA^-^/CD127^+^/CCR7^-^) and exhausted (CD4^+^Tex; PD-1^+^/TIM3^+^) CD4 T cell populations (Figure 2D). Upon *in-vitro* stimulation, the expression of CXCR3 on the cell surface (Mann-Whitney U test, p= <0.01), intracellular IL-4 levels (p= 0.04) and IL-4, IL- 6 in combination were found to be significantly lower in the CD4 T of PVS cohort (p= <0.01), with no differences were observed in IFN*γ* & TNF⍺ levels (Figure 2E). Significant increases in intracellular TNF⍺ levels (p= <0.01) with non-significant increases in IFN*γ* in the stimulated CD8 T cells were observed in PVS cohort (Figure 2F). Only a total of 32.23% of the variability in intracellular TNF⍺ could be explained by IFN*γ* levels in the CD8^+^ T cell populations (R^2^= 0.32; Figure 2G).

In the subgroup analyses, no differences in proportions of DN B cell subpopulations were observed (Figure S2D). Proportions of CD4^+^ CD45RA^+^ effector memory T cells (CD4^+^ T_EMRA_; CD45RA^+^/CD127^-^/CCR7^-^; p= 0.02) and rnTregs (p= 0.03) were both observed to be significantly lower in PVS+I compared to the control+I subgroup (Figure S2E). Proportions of CXCR3 expressing stimulated CD4 T cells was much lower in PVS+I cases (p= <0.01) and the proportions of both IL-4^+^ (p= 0.04), and IL-4^+^/IL-6^+^ (p=< 0.01) double-positive cells were also lower compared to the controls (Figure S2E). Higher proportion of CD8^+^ Tcm cells was retained in the PVS+I subgroup compared to the control+I group (Figure S2F; p= 0.02). No differences were observed in immune cell populations between the infection-negative cases (PVS-I) and controls (Controls-I).

### Lower levels of spike-specific antibody responses in PVS

Given the differences in the number of vaccine doses received between participants in the PVS cohort and the control group, we compared spike-specific immunoglobulin G (IgG) levels in relation to the number of vaccine doses administered. Correlation analyses revealed a significant positive correlation between the number of vaccine doses and plasma anti-S IgG levels (Spearman’s Rank Correlation Coefficient, ⍴ = 0.85, p= <0.01) as well as anti-RBD IgG levels (⍴ = 0.83, p= <0.01) in the PVS-I subgroup. In the PVS+I subgroup, only anti-S IgG levels showed a significant correlation (⍴ = 0.55, p = 0.01) with the number of doses (Figure 3A). Next, correlation analyses were performed to assess the relationships between plasma anti-S, anti-RBD, and anti-N IgG levels with the number of days post last vaccination among the four groups. No significant changes in anti-S and anti-RBD antibody levels were observed with increasing days since vaccination in the control group, regardless of infection history, and in the PVS+I subgroup (Fig 3B). In contrast, significant negative correlations were found in the PVS-I subgroup between the number of days post-vaccination and both anti-S (⍴ = −0.87, p= <0.01) and anti-RBD (⍴ = −0.83, p= <0.01) IgG levels, indicating a decline in these antibodies over time (Fig. 3B). Additionally, as expected, no correlations were observed between anti-N IgG levels and days post vaccination across the infection-positive subgroups.

**Figure 3:**
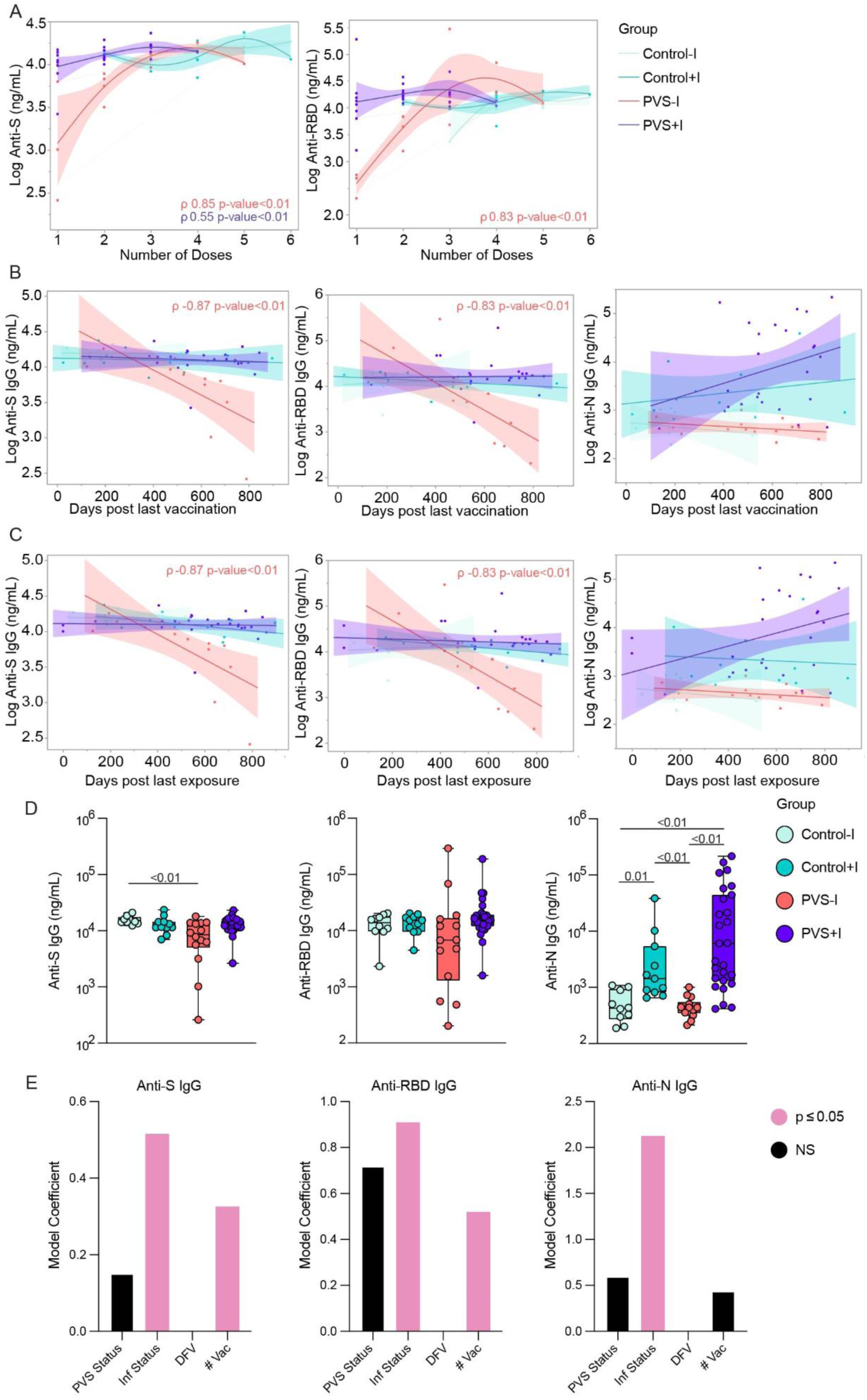
Plasma reactivity to SARS-CoV-2 antigens. **A.** Correlation comparisons of virus-specific ancestral anti-S and anti-RBD IgG levels by number of COVID vaccine doses. **B.** Correlation and linear regression comparisons of virus-specific ancestral anti-S, anti-RBD and Anti-N IgG levels by days post last vaccination. **C.** Correlation and linear regression comparisons of virus-specific ancestral anti-S, anti-RBD and Anti-N IgG levels by days post last exposure. Regression lines are shown colored by groups Control-I, Control+I, PVS-I, and PVS+I as indicated in the figure legend. Spearman’s ρ coefficients and linear regression significance are colored; accordingly, shading represents 95% confidence interval. **D.** Plasma reactivity to ancestral S, RBD, and N proteins measured by ELISA are shown by groups Control-I, Control+I, PVS-I, and PVS+I. Significance of difference in group median values was assessed using Kruskal– Wallis with Benjamini–Hochberg false-discovery rate (FDR) correction for multiple comparisons. The central lines indicate the group median values, and the whiskers show the 95% CI estimates. **E.** Generalized linear model analysis for virus-specific ancestral anti-S, anti-RBD and Anti-N IgG levels. Model predictors are indicated on the x axis and include days from vaccination (DFV) among others. Predictors with p ≤ 0.05 are highlighted in pink to indicate significance, while non-significant predictors are displayed in black. Detailed model results are shown in table S3.

The next step was to evaluate if the most recent exposure to SARS-CoV-2 or vaccination correlated with the observed differences in waning patterns. No significant changes in anti-S, anti-RBD and anti-N antibody levels were observed with an increase in the number of days from self-reported viral infection dates among the Control+I and PVS+I subgroups (Figure 3C). In addition, the plasma titers of anti-S IgG were significantly lower among the PVS-I cases compared to the Control-I subgroup (p= <0.01) (Figure 3D). However, no differences were observed in the anti-RBD IgG levels across the four subgroups (Figure 3D). As expected, the uninfected PVS-I and the Control-I subgroups had much lower anti-N IgG levels as detected by in-house ELISAs (Figure 3D). To further account for variations in vaccine doses and infection, we developed linear models. Those models indicated that both prior SARS-CoV-2 infection and the number of vaccine doses were significantly associated with higher levels of anti-RBD and anti-S IgG (Figure 3E, Table S3).

### Serological evidence of recent EBV reactivation in PVS

Many human pathogens are ubiquitous, opportunistic, and capable of establishing lifelong infections with alternate latency and reactivation cycles^26^. These cycles can be triggered by physiological perturbations and can contribute to systemic inflammation^27^. Therefore, we used serum epitope repertoire analysis (SERA) to evaluate seropositivity against a range of pathogens, including five bacterial, seven parasitic,14 viral and one fungal species. On performing two group analyses, no significant differences were observed for all pathogens, indicating similar levels of prior exposure. (Fig. 4A).

Moreover, the seropositivity for each pathogen did not significantly differ from seropositivity in 3448 healthy controls collected before the COVID-19 pandemic (Figure 4A). Given the high seropositivity rates for herpesviruses, we further analyzed the seropositivity patterns in combination, for cytomegalovirus (CMV), Epstein-Barr Virus (EBV), Herpes Simplex Virus Type 1 (HSV-1) and Herpes Simplex Virus Type 2 (Figure 4B). Significant differences were observed between cases and controls (Mann-Whitney U test, p = 0.01; Figure 4C), where the participants with PVS had higher prevalence of EBV and HSV coinfection, and lower prevalence of EBV and CMV coinfection. There are reports of similarities in symptom phenotypes between PVS and long COVID, as well as evidence of EBV reactivation in long COVID cases, including elevated antibodies against EBV surface protein gp42^22, 28^. Therefore, we further investigated the prevalence of antibodies against EBV gp42 and identified significantly elevated antibodies in the plasma of PVS participants compared with controls (Kruskal-Wallis test, p = <0.01, Figure 4D, E). As an orthogonal validation, we tested the distribution of linear peptide reactivities across the EBV proteome. Greater reactivities to two peptides corresponding to two envelope glycoproteins necessary for B cell infection, gp42 and gp350 were observed. For the gp42 protein, the antibody reactivity to peptide ([VI]XLPHW) was significantly higher among the PVS participants irrespective of the SARS-CoV-2 infection status (Mann Whitney U test, p= <0.01; Figure 4F) and across the four subgroups (Kruskal-Wallis test, p= 0.02; Figure 4G). Greater reactivities were also observed for the gp350 peptide (KXRX[RQ]WXF) among the PVS participants compared to controls (Kruskal-Wallis test, p= 0.04; fig 4J) and across the four subgroups (Kruskal-Wallis test, p= 0.03; Figure S3C). Further, anti-gp42 ([VI]XLPHW) reactivity by SERA significantly correlated with anti-gp42 ELISA measurements thus validating the finding (R = 0.37, p= <0.01; Figure 4I). We also mapped this motif onto available structures of gp42 complexed with EBV gH/gL (PDB: 5T1D), demonstrating its location close to the transmembrane (TM) domain of gp42 and surface-exposed (Figure 4H). Study participants with greater antibody reactivity to gp42 as assessed by ELISA also exhibited higher percentages of TNFα-producing CD8+ T-cells (R = 0.47, p= <0.01, Figure 4K). This correlation was not observed for IL-4, IL-6 double-positive CD4 T cells (Figure S3D) as was previously reported for long COVID^22^.

**Figure 4:**
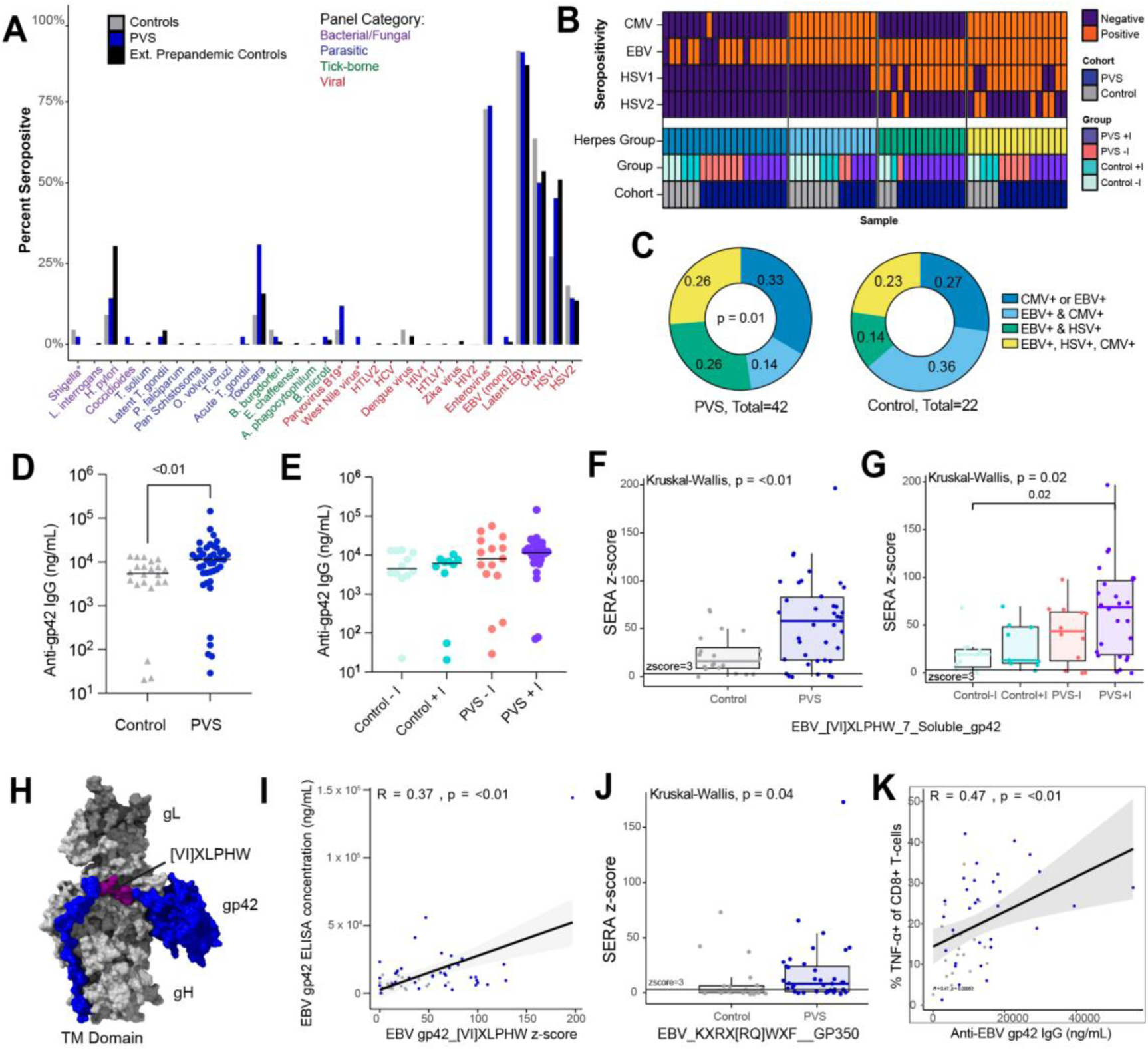
Elevated responses to Epstein Barr Virus in PVS patients. **A.** Proportion of each group (PVS: n = 42, control: n = 22, pre-pandemic healthy control: n = 3448) seropositive for each of 31 common pathogen panels as determined by SERA, grouped by pathogen-type. Statistical significance determined by Fisher’s exact test corrected with FDR (Benjamini Hochberg). Star indicates panels for which pre-pandemic healthy controls were not analyzed. **B.** Heatmap showing supervised clustering of SERA-determined seropositivity to EBV, CMV, HSV-1, and HSV-2 across samples. Clusters were named for their herpesvirus dominance and are labeled accordingly. **C.** Herpes seropositivity composition for each cohort. Significance of relative enrichment for each cluster was calculated using Chi-square test of observed composition vs. expected composition. **D, E** Plasma reactivity to EBV gp42 protein measured by ELISA shown by cohort, PVS and Control **(D)** and by groups Control-I, Control+I, PVS-I, and PVS+I **(E)**. **F.** SERA-derived *z* scores for the gp42 motif [VI]XLPHW among EBV-seropositive individuals only, plotted by cohort, *n* = 20 (Control), *n* = 38 (PVS) **(F)** and group, *n* = 11 (Control-I), n = 9 (Control+I), *n* = 12 (PVS-I), *n* = 26 (PVS+I) **(G)**. The dashed line represents the *z*-score threshold for epitope positivity defined by SERA. **H.** Three-dimensional mapping of the PVS-enriched linear peptide sequence [VI]XLPHW (purple) onto EBV gp42 (blue) in a complex with gH (light grey) and gL (dark grey) (PDB: 5T1D). **I.** Relationship between EBV gp42 [VI]XLPHW SERA *z* score and plasma concentration of anti-gp42 IgG. **J.** SERA-derived *z* scores for the gp350 motif KXRX[RQ]WXF among EBV-seropositive individuals only, plotted by cohort. The dashed line represents the *z*-score threshold for epitope positivity defined by SERA. **K.** The relationship between plasma concentration of IgG against EBV gp42 and the percentage of TNFα CD8+ T cells (of total CD8+ T cells). For all box plots, the central lines indicate the group median values, the top and bottom lines indicate the 75th and 25th percentiles, respectively, the whiskers represent 1.5× the interquartile range. Each dot represents one individual. Statistical significance of the difference in median values was determined using Kruskal–Wallis tests with Post hoc Dunn’s test and Bonferroni–Holm’s method to adjust for multiple comparisons. Correlation was assessed using Spearman’s correlation. The black line shows linear regression, and shading shows the 95% CIs.

### Participants with PVS have a distinct set of autoantibodies

To evaluate differences in immunoglobulin isotypes and IgG subtypes in the plasma, Luminex assays were performed. No significant differences were observed between the PVS cohort and the controls (Figure S4A). Next, to determine the presence of autoantibodies in PVS, we screened for reactivities across a range of 120 known autoantigens using microarrays for three different immunoglobulin isotypes, IgM, IgG, and IgA. We observed significant increases in IgM reactivities against 65 antigens, IgG reactivity against 1 antigen and IgA reactivities against 39 antigens in PVS compared to controls after multiple testing corrections (Table S4). Among these antigens, two showed log_2_fold change of greater than 2: anti-nucleosome IgM (Mann-Whitney U test, p= <0.01) and anti-AQP4 IgA (p= <0.01) (Figure S4B). Conversely, control participants exhibited higher reactivities against a total of 21 antigens, 18 of which were of the IgG isotype and five were of IgA isotype with two common antigens between the two isotypes (Table S3). Among these autoantibodies, anti-histone H1 IgG differed by greater than log_2_fold change (p= <0.01, Figure S4B). Infection-positive subgroups had a higher number of reactivity differences between cases and controls (Figure S4C). Among the PVS-I participants, anti-calprotectin/S100 IgM, anti-genomic DNA IgA and anti-ssDNA IgA reactivities were significantly higher while anti-histone H3 IgG, anti-MBP IgA, and anti-PR3 IgA reactivities were higher among the controls-I (Figure S4C, Table S5).

### Circulating hormones and immune modulators in PVS

Two group analyses of circulating hormones and immune modulators revealed significantly lower levels of fetuin A26 and neurotensin (Mann-Whitney U tests, p= 0.01 and p= 0.03; Figure S5A) in participants with PVS with fold differences of 1.3 and 1.9 respectively. Additionally, four group analysis was performed to evaluate the impact of infection on PVS. Given the smaller number of samples in the four group analyses, each panel of analytes was independently evaluated. Significantly lower levels of circulating fetuin A36, and neurotensin were also observed among participants with PVS with a history of SARS-CoV-2 infection compared to convalescent participants (p= 0.01 for both analytes; Figures S5B-C). No differences were observed for other factors across the subgroups except for β endorphin which was significantly lower in PVS+I compared to the control+I group (p= 0.01; Table S7) without any significant differences in the two group analyses. No differences were observed in the uninfected subgroups.

### Increase in circulating SARS-CoV-2 Spike protein in participants with PVS

It has been reported that the BNT162b2 or mRNA-1273 derived S proteins circulate in the plasma of those vaccinated as early as one day after the vaccine and interactions of the circulating protein^16^. Hence, we next sought to investigate whether the S1 subunit of the SARS-CoV-2 S protein could be detected in the plasma. For this, we used an anti-S1 Successive Proximity Extension Amplification Reaction (SPEAR) immunoassay. This method can detect S1 levels as low as 5.64 fM. We conducted a one-sided Kolmogorov–Smirnov test with 1000 permutations to see if the participants with PVS had higher circulating S1. The results indicated that participants with PVS had significantly higher circulating S1 levels compared with the control group (p = 0.01). However, circulating S1 was found in only a subset of participants with PVS at varying concentrations while the control group mostly exhibited a bimodal distribution of zero and non-zero values (Fig. 5A, Table S2). Detectable S1 was found in participants’ plasma ranging from 26 to 709 days from the most recent known exposure (Figure 5B). To fully account for the width of this dataset, we included all non-detectable values in the analysis and applied a generalized regression model accounting for zero-inflation. We found that both PVS-I and PVS+I groups displayed significantly elevated S1 levels than the Control-I group (p= <0.01 and p= 0.02, respectively) (Figure 5C).

**Figure 5:**
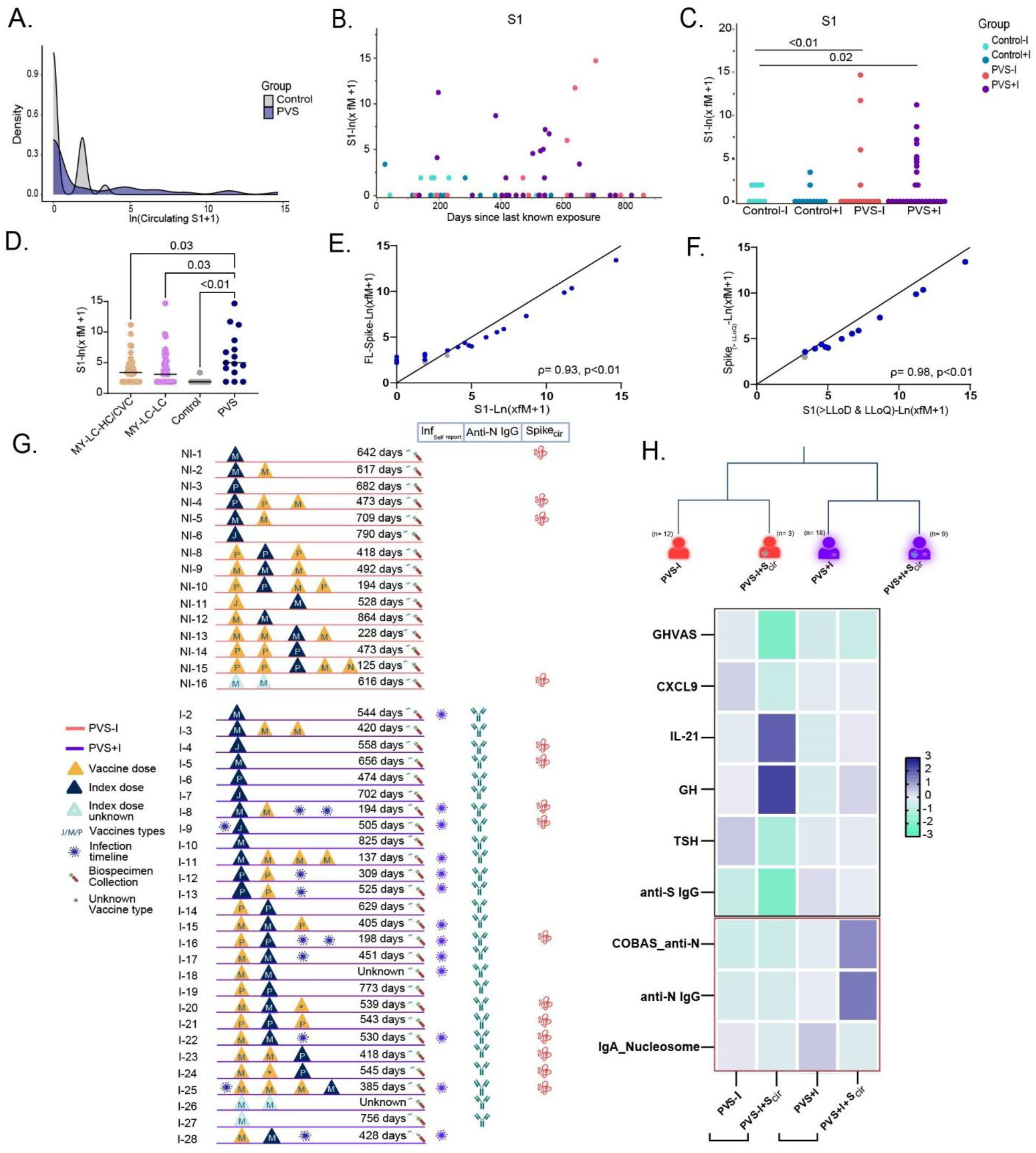
Circulating SARS-CoV-2 Spike protein. **A.** Density plots describing the distribution of circulating S1 levels across controls (n= 22) and PVS (n= 42) measured by SPEAR assays. **B.** Levels of circulating S1 in plasma days post last known self-reported exposure. **C.** Circulating S1 levels measured by SPEAR assay are shown across groups Control-I, Control+I, PVS-I, and PVS+I. A parametric test incorporating a zero-inflated Poisson model was used to account for the excess zeros in the data. **D.** Circulating S1 antigen levels above LLoD across cohort groups, MY-LC-HC/CVC (n= 41), MY-LC-LC (n=45), Control (n= 7), PVS (n= 15). **E.** Correlation between circulating Spike protein assays using antibodies for the S1 subdomains and S1& S2 subdomains (full length Spike). **F.** Correlation between circulating Spike protein assays among samples with values above LLoD and LLoQ. Correlations were assessed using Spearman’s rank correlation. **G.** A participant-specific graphic representation illustrating key events including vaccination, infection, sample collection and the presence or absence of anti-N antibodies and circulation spike protein. Each participant is represented by a single horizontal line and each vaccination event is marked by triangles, and the index doses are highlighted in blue. The number of days between latest exposure and biospecimen collection is also indicated. The abbreviations for the vaccine types are: J for Jcovden (Johnson & Johnson), M for Spikevax (Moderna), and P for Comirnaty (Pfizer-BioNTech). **H.** A classification tree of PVS participants based on infection status with or without detectable S1 in circulation and a heatmap of distinct demographic and immunological variables that differentiate PVS within the infected and uninfected subgroups based on Mann-Whitney U tests.

Given the similarities between PVS and long COVID symptoms, one hypothesis in the literature is that shared exposure to the S protein may play a role and several groups have independently reported the presence of circulating S1 & full-length S in long COVID using various detection methods^16,29^. To further investigate this circulating S1 positivity percentages and levels in the LISTEN cohort subgroups were compared with an external cohort of healthy, convalescent controls and LC participants (MY-LC cohort). This external cohort, collected from Mount Sinai clinics, included 134 healthy/convalescent controls and 134 long COVID participants, which were all assayed together with the LISTEN cohort biospecimens.

Among the MY-LC healthy [HC(n= 62); no reported SARS-CoV-2 infection], convalescent controls [CVC(n=72); with reported SARS-CoV-2 infection], and long COVID participants, detectable S1 was observed in 30.6% of control participants (41/134; HC= 12.9%; CVC= 22.2%) and 33.6% (45/134) of individuals with long COVID, with mean S1-ln(x_fM_ +1) values of 3.72 and 3.85 respectively among those above the LLoD. These figures were comparable to the percentages observed among the LISTEN controls (31.8%) and PVS (35.7%) groups. S1 levels were moderately elevated in the MY-LC control group compared to the PVS control group (p=0.06), potentially reflecting differences in exposure timing or SARS-CoV-2 variant of concern (VOC). Despite this, the PVS group demonstrated significantly higher S1 levels compared to both control cohorts (LISTEN-control: p<0.01; MY-LC control: p=0.03; mean S1-ln(x_fM_ +1) = 6.24) (Fig. 5D, Table S8).

To further validate the findings and to investigate whether the presence of S1 reflects the presence of full-length S protein among the LISTEN participants, we next conducted a full-length S SPEAR assay. The calculated values for the LLoD and the LLoQ were 1.81 and 8.24 fM, respectively. The full-length S SPEAR assays showed a significant correlation between S and S1 across all samples (Fig. 5E), as well as for values above the LLoD and LLoQ (Fig. 5F, Table S2). Thus, based on SPEAR assays, the individuals with PVS exhibited elevated levels of circulating full-length S compared to healthy controls.

### Immune signatures in PVS subgroups based on the presence of circulating S1 protein

To gain a clearer understanding of the variability of circulating S1 protein levels, we first compiled a structured timeline that summarizes the self-reported infection dates, vaccine numbers (including types and administration dates), and the number of days between the latest known exposure and the collection of biospecimens. This timeline was organized for both the PVS-I and PVS+I groups (Figure 5G). Notably, we observed that the highest levels of detectable S1 in the PVS-I group were the furthest away from the last known exposure and ranging between greater than 600-700 days (NI-1 & NI-5; Figure 5G). This suggested that prolonged antigen persistence might be associated with PVS in a subgroup of patients. Further, most of the PVS+I group participants experienced breakthrough SARS-CoV-2 infections with the exception of two cases, indicating that PVS symptoms started prior to infection (Figure. 5G).

Given the possible heterogeneity in immunological trajectories leading up to PVS and the lack of adequate sample numbers in each PVS subgroup, we next took a more descriptive approach to look for peripheral immune signatures stratified based on their infection status and detectible S1 above SPEAR assay’s lower limit of quantitation. In order to begin with a valid method of selection despite the small sample sizes, non-parametric Mann-Whitney tests were implemented without multiple testing corrections to look for differences in distributions of 547 independent variables including GHVAS scores, circulating modulator levels, anti-SARS-CoV-2 antibody titers and autoantibody scores within both the PVS-I and PVS+I subgroups. Variables showing significant differences were further filtered based on greater than 1.5 fold changes to identify the distinct determinants associated with each of the four trajectories.

Among other factors, the infection-naïve PVS participants with quantifiable S1 had lower GHVAS scores indicative of poorer general health (GHVAS) and lower anti-S IgG titers, whereas higher circulating IL-7 and IL-21 levels were detected compared to other groups (Figure. 5H). Elevated growth hormone levels alongside low TSH levels were also observed among the PVS-I participants with S1 protein in circulation. By contrast, among the infection-positive subgroups, participants with circulating S1 were observed to have higher anti-N antibody titers based on the clinical COBAS assays and in-house ELISA indicative of the contribution of infection history. Moreover, anti-nucleosome IgA levels were higher among those in the PVS+I subgroup without detectable S1 (Figure. 5H).

### Machine Learning-based identification of peripheral immune signatures of PVS

To establish a combined global immune signature for persistent symptoms following COVID-19 vaccination, we built machine learning models to predict PVS outcomes. The goal was to identify prominent features that could effectively distinguish PVS from the controls in a parsimonious manner. Given that autoantibodies are also common in the general population at low levels, we chose to exclude them from this analysis in the absence of further validation^30^. Additionally, we excluded any variables with greater than 20% missing values for either the PVS or control groups and SERA variables because most of the dataset lacked a significant number of values above LLoD. A total of 193 variables were included.

Weighted Gene Co-Expression Network Analysis (WCGNA)^31^ was applied to this set to find groups of highly correlated variables (Figure S6B). The final feature set was then created by taking all variables that did not form a tight cluster (141, Module 1) and the eigengene, or first principal component, of every other set of variables (Modules 2-6). This gave us a total of 146 features. Next, we performed classification utilizing Least Absolute Shrinkage and Selection Operator (LASSO) with nested cross-validation. We achieved an overall model accuracy of 78.1% on the validation folds (per-fold range 62.5% - 100%; Figure S6A), and an AUC of 0.80 (per-fold range 0.67-1.00, 95% CI = 0.67-0.92; Figures 6A and S6A). A permutation test further concluded that this performance was significantly above random (p = 0.02; Figure S6A). Segregating test data per-fold by infection status yielded accuracy on the infected population of 86.5% and the uninfected population of 73.3%. (Figure S6A) Per-class accuracy showed some divergence, with PVS accuracy of 85.7% vs. control accuracy of 63.6%. This was due to a subset of the controls clustering primarily with PVS samples, making control classification more difficult (Figure 6B).

**Figure 6:**
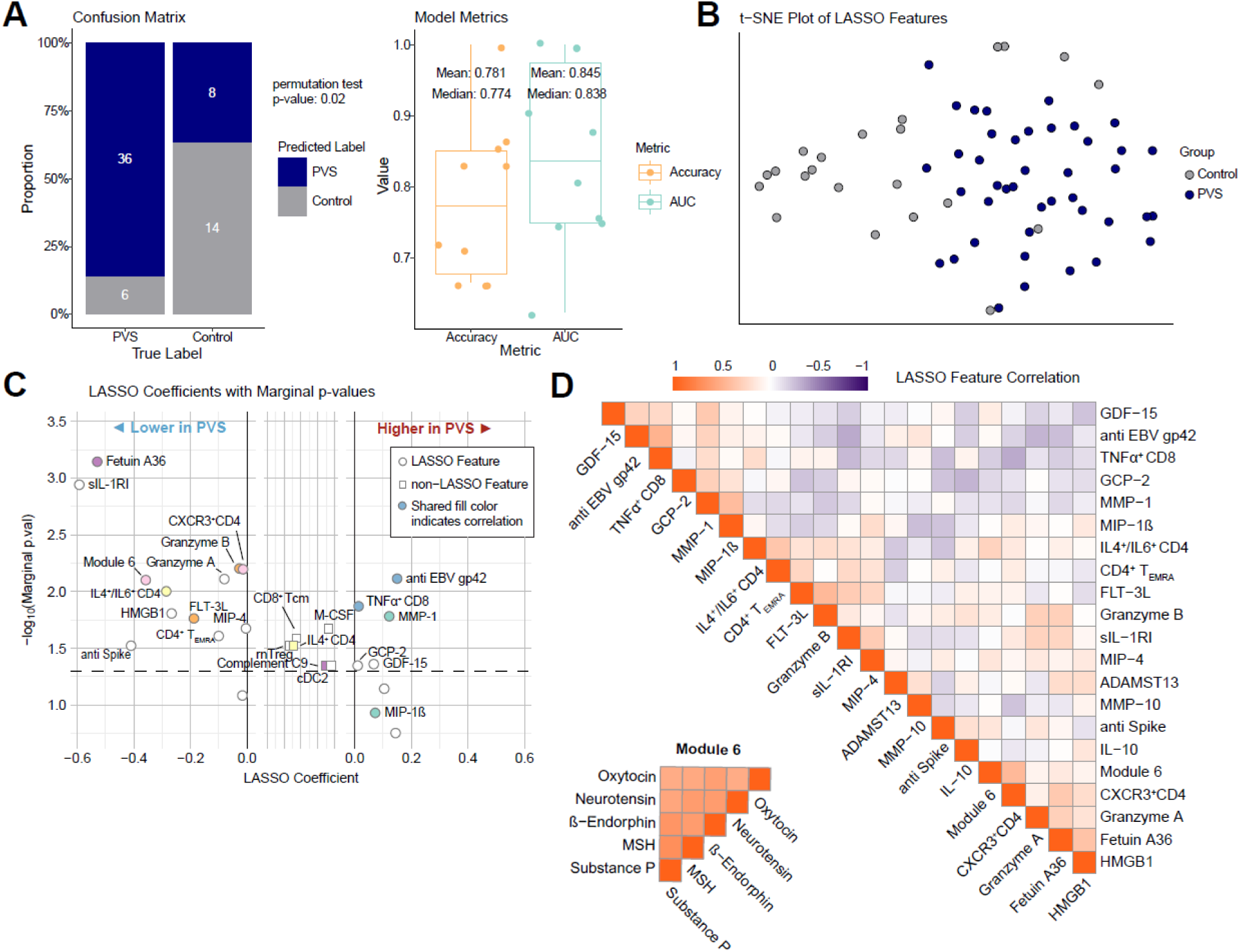
Machine Learning Results and Prominent Feature Identification. **A.** Confusion matrix inspired barplot describing actual and predicted labels for PVS and control with a classification threshold of 0.65. The p-value from a permutation test is also displayed to motivate model legitimacy. Boxplot displaying accuracy and AUC for each outer fold of nested-cross validation with both inner and outer loops set to 10. Mean and median accuracy and AUC stats are also shown. **B.** t-SNE plot showing a two-dimensional similarity-based representation of the LASSO variable space. **C.** Scatter plot containing all variables identified through either LASSO or marginal logistic regression models. Features only identified by marginal test, and not by LASSO are displayed in the center panel with coefficient of 0. Points with a shared color display Pearson correlation > 0.4. **D.** Correlation heatmaps of LASSO-selected and WGCNA module six features, with features clustered through hierarchical clustering.

The LASSO model selected 21 features using all data, consisting of CD4 T cell populations, immune modulators, neuropeptides, and antibodies (Figure 6C). Among the features selected, there were several negatively associated with PVS. These included circulating factors sIL-1R1, fetuin A36, granzyme A and B, FLT−3L and HMGB1, and subsets of circulating CD4 T cell populations (CXCR3^+^ CD4 T cells CD4^+^ T_EMRA_ cells, and IL-4^+^/IL-6^+^ CD4 T cells). Multiple hormones and neuropeptides synthesized by the hypothalamus, pituitary glands, and the peripheral nerves and involved in nociception and stress responses such as oxytocin, neurotensin, ꞵ endorphin, melanocyte-stimulating hormones (MSH), and substance P were also negatively associated with PVS and formed a single module (Module 6) (Figure 6D). The features that were positively associated with PVS were anti-EBV gp42 IgG titers, MMP1 levels, and TNFɑ^+^ CD8 T cells. We observed that no single variable or small subset of variables had a particularly strong differentiating power.

## DISCUSSION

In this study, we examined symptoms and circulating immune factors and cell types associated with chronic illness following COVID-19 vaccination. Post-acute conditions following COVID-19 vaccination have been reported for multiple vaccine platforms including mRNA and adenoviral-vectored vaccines^6,7,8,9^. We observed that the general health status of the PVS participants was far below the general US population average^32^ based on the GHVAS scores. The patient-reported outcome scores from the PROMIS29 domains were also indicative of lower quality of life. A large fraction of individuals reported the onset of symptoms to be as early as within one day of COVID-19 vaccination. Compared with controls, participants with PVS had reduced CD4^+^ T cell subsets in circulation (both Th1 and Th2) and an increased percentage of TNFα^+^ CD8 T cells. Among cell populations of myeloid origin, cDC2 cells were reduced, and non-classical monocytes were elevated among PVS participants. Lower S-specific IgG levels were observed in PVS mainly due to the limited vaccine doses received. Additionally, serological evidence for recent EBV reactivation was also observed. Using machine learning approaches, we further identified a set of 21 core predictive features of PVS status within the LISTEN PVS cohort with potential for further validation and biomarker identification. Most notably, we found elevated levels of spike (S1 and full-length S) in circulation up to 709 days after vaccination among a subset with PVS, even in those with no evidence of detectable SARS-CoV-2 infection.

To date, only a few studies have investigated the immunological mechanisms associated with PVS ^11,12, 33^, and no consensus definition of this syndrome exists ^10, 34^. Previous studies on PVS have found the presence of elevated levels of inflammatory cytokines such as CCL5, IL-6, and IL-8; IgG subclass imbalances, high angiotensin II type 1 receptor antibodies (AT1R), and the presence of spike S1 in non-classical monocytes, among others^11,12,33^. In the LISTEN PVS cohort, we did not find evidence of elevation in inflammatory cytokines or IgG subclass imbalances. This difference may be due to the heterogeneity of the cohorts studied, vaccine types or the time from vaccination.

The demographics at risk of developing PVS and symptom manifestations are similar to those of long COVID ^10, 35,36,37^. Whether this reflects overlapping underlying mechanisms such as persistent S protein remains to be determined. Circulating S1 antigen has been detected in mRNA-1273 vaccine recipients without a prior history of viral infection within an average of five days after the first injection and becomes undetectable by day 14^16^. By contrast, in our study, significantly elevated levels of circulating S1 and S were observed in a subset of PVS participants both in the infection-naive and infection-positive groups up to 709 days post-exposure. This is in line with the findings of S1 persistence in monocytes in people with PVS^12^. Circulating full-length S has also been detected in cases of post-vaccination myocarditis^38^. Given the striking similarities between long COVID and PVS symptoms, there has been speculation regarding the potential causal role of the persistent presence of spike protein^39^ driving the chronic symptoms. Additionally, a recent study has shown spike protein binding to fibrin resulting in inflammation *ex vivo* and neuropathy in animal experiments^40^. S1 subunit is sufficient to cause formation of trypsin-resistant fibrin clots when added to plasma from healthy individuals^41^. The persistent presence of S1 and the full-length spike protein across multiple long COVID cohorts lends further support to this hypothesis^42,43,44,45^. Additionally, our results using the S1 SPEAR assays indicate higher percentages of individuals with S1 antigen persistence among both MY-LC controls and the long COVID group compared to other studies despite reporting mild acute phase symptoms reported by these participants ^44,45,43^. This may be attributed to variations in assay sensitivity or variations in vaccine doses and re-infection rates across the cohorts. Despite higher antigen persistence rates, the PVS participants with detectable S1 had higher mean circulating S1 levels compared to the LC participants. In our PVS-I group, anti-S antibody levels were lower in those with circulating S1. Why persistent spike antigen fails to elicit an antibody response, and what the source of persistent spike in circulation is, requires further investigation.

Immunophenotyping of circulating PBMCs from participants with PVS revealed lower levels of circulating CD4^+^ Tem, CXCR3 expressing CD4, as well as IL-4^+^/IL-6^+^ double positive CD4 T cell populations and higher TNFα secreting CD8 cell populations. This is in contrast to our observations of higher levels of IL-4^+^/IL-6^+^ CD4 T cell populations in the long COVID cohort^22^. Elevated levels of anti-S IgG have been observed in long COVID patients, possibly reflecting persistent S protein^22,42^. By contrast, within the PVS-I subgroup, the lower levels of anti-S antibodies were associated with a reduced number of vaccinations. Moreover, PVS participants in this study did not exhibit decreased circulating cortisol levels or increased fetuin A36 levels, as reported for long COVID^22,46^.

While our panel of autoantigens did not include any G protein-coupled receptors included in the Semmler et al study^11^, among those identified to be elevated in PVS in this study, namely, anti-nucleosome IgM and anti-AQP4 IgA require further investigation. Higher monomeric IgM has been reported in autoimmune disease patients and circulating nucleosomes have also been shown to trigger cGAS (cGMP-AMP synthase) immune responses^47,48^. Along similar lines, anti-AQP4 IgG is most commonly associated with neuromyelitis optica spectrum disorder (NMOSD), however there are not any reports on IgA isotype^49^. In addition, similar to what has been reported in long COVID, elevated antibody responses against EBV lytic antigen were detected among seropositive participants with PVS, suggesting recent reactivation, ^22,28^.

This study has several limitations. Our small sample size could have affected the robustness of the machine learning approaches and prediction of specific immune features in PVS. Due to the limited sample size, we might have failed to capture small but potentially important immune features associated with PVS. Analysis of autoimmune antibody reactivity was restricted to antigens reported in other autoimmune diseases, limiting the discovery of a broader range of autoantibodies. While we used two independent approaches to ascertain previous infection with SARS-CoV-2, negative results cannot definitively preclude prior infection that occurred in the distant past. Other limitations include the lack of analysis of the host genetics that might account for PVS susceptibility, or any other conditions, such as non-prescription drugs or asymptomatic infection with other pathogens that were not tested in our analysis might have predisposed an individual to develop chronic illness following COVID-19 vaccination. While we observed elevated levels of S1 among those with PVS compared to LC, additional studies with matched patient demographic profiles are necessary to determine whether this represents genuine differences or is simply a result of random variation. Finally, we do not know whether our findings extend beyond COVID-19 vaccination since we did not include PVS following other vaccines.

In summary, by revealing distinct immunological features of PVS, this study helped generate hypotheses regarding the underlying pathobiology of this condition. Understanding such mechanisms will help improve the overall safety profile of COVID-19 vaccines and support public health strategies that maximize vaccine efficacy while minimizing adverse effects. However, this study is early-stage and requires replication and validation. We emphasize the critical task of discerning between meaningful results and random fluctuations in the data. Future work is essential to elucidate these relationships. As the global community continues to navigate the challenges of COVID-19 and long COVID, a deeper understanding of vaccine-related immune responses will be essential in refining vaccination practices and ensuring their long-term success.

## Data Availability

The flow data repository ID is FR-FCM-Z8FZ for all the raw .fcs files generated for flow cytometry analyses at the Flow Repository platform. Custom codes used for computational analyses will be made available by authors upon reasonable request.

## RESOURCE AVAILABILITY

The flow data repository ID is FR-FCM-Z8FZ for all the raw .fcs files generated for flow cytometry analyses at the Flow Repository platform ^50^. Custom codes used for computational analyses will be made available by authors upon reasonable request.

## ACKNOWLEDGEMENTS

We thank all the participants from the LISTEN study who contributed their time, effort and precious biospecimens for this study. We thank the Yale Center for Clinical Investigation (YCCI) coordinators Geisa Wilkins, Iris Dias and Yvette Strong for their support. Victoria Bastos’ fellowship was supported by CAPES Foundation, Ministry of Education, Brazil. This study was funded in part by the Howard Hughes Medical Institute Collaborative COVID-19 Initiative.

## AUTHOR CONTRIBUTIONS

Experimental conceptualization, methodology and data visualization were performed by B.B, P.L.,V.B., V.S.M., A.T., K.W. W.B.H, K.G., K.K. and J.R. Formal analysis was conducted by B.B, P.L.,V.B., V.S.M., A.T., K.W. W.B.H and K.G. Resources were provided by H.M.K and A.I. Review of survey responses and electronic health records were performed by B.B. Sample collection, processing and biospecimen validation were performed by B.B., T.J.T, P.B. and C.G., The original draft was written by B.B. and A.I. Review and editing were performed by B.B., P.L.,V.B., V.S.M., A.T., K.W. W.B.H, K.G., K.K., J.R., D.H., B.D., L.G, H.M.K, and A.I. Data curation was performed by B.B., W.B.H, F.W. L.G. M.S., and H.M.K. and A.I. supervised the study. Funding was acquired by A.I.

## MATERIALS AND METHODS

### Ethics Statement and Study Design

A retrospective, decentralized, exploratory case-control study was conducted on vaccinated participants ≥ 18 years of age enrolled in the LISTEN study which recruited individuals from the Hugo Health (Guilford, CT, USA) Kindred community ^51^. This study was approved by the Yale University Institutional Review Board on April 1, 2022 (HIC# 2000032207) and conforms to the Declaration of Helsinki and STROBE reporting guidelines^52,53^. Informed consent was provided by participants electronically. Only those residing in contiguous US states were included in this arm of the study. Each participant was assigned a unique identifier as part of the de-identification protocol managed by the study coordinator. These identifiers were kept confidential and were not accessible to anyone outside the research team.

A subset of the MY-LC cohort recruited from within the Mount Sinai Healthcare System and the Centre for Post COVID Care at Mount Sinai Hospital which included healthy and convalescent controls and long COVID participants were included for validation of one of the assays. The biospecimens were collected between 2021 and 2023 as approved by the Mount Sinai Program for the Protection of Human Subjects (#20-01758).The recruitment for the MY-LC cohort has been elaborately described in the Klein et al study ^22^.

### Inclusion and Exclusion Criteria

Given the exploratory nature of this study, no power analysis was done. Only US residents were included. Cases were defined as participants who self-reported having PVS after administration of the primary or booster series of COVID-19 vaccine and controls were vaccinated individuals who reported no adverse events. The cohort consisted of both individuals with self-reported history of SARS-CoV-2 infection or otherwise. Participants with a medical history of arthritis, asthma, autoimmune disease, blood clots, cancer, cerebrovascular conditions, chronic lung disease (including emphysema, chronic bronchitis, chronic obstructive pulmonary disease), cystic fibrosis, diabetes, Ehlers Danlos Syndrome, myocardial infarction, heart conditions, heart failure, history of organ transplant, immunocompromised state, kidney disease, liver disease, Lyme disease, mast cell activation syndrome, myalgic encephalomyelitis/chronic fatigue syndrome, neurologic conditions, postural orthostatic hypotension, spinal disorder(s), tremors/Internal vibrations, hyperthyroidism, hormonal migraines, schizophrenia spectrum disorders, bipolar disorders and somatic symptoms before 2020 were excluded. Participants who self-reported having both long COVID and PVS were also excluded from this study. Among the selected cases, those with GHVAS scores of ≤75 at the time of screening were included, while among the controls, only those with GHVAS scores of ≥80 or general health of very good/excellent were included.

Electronic health records, shared by participants via Hugo Health were accessed to obtain a participant-wise list of ongoing medications to exclude participants on immunosuppressants.

### Survey Instruments

Members of the Hugo Health Kindred community were offered a set of specialized baseline survey instruments. The first questionnaire was geared towards gathering information on demographic variables such as age, biological sex, ethnicity as well as self-reported medical conditions such as long COVID and PVS. The subsequent surveys were on self-reported SARS-CoV-2 infection, vaccination profiles and medical history, comorbidities, PVS symptoms ^10^. These survey responses were analyzed, and a subset of the participants were included in this arm of the study based on the inclusion and exclusion criteria mentioned above.

At the time of biospecimen collection, the participants were asked to rate their general health status using the following options: excellent, very good, good, fair, poor and do not know and a general health visual analogue scale (GHVAS) having a numerical value between 0 to 100, where higher scores represented better health. These responses were recorded by ExamOne phlebotomists on LISTEN-ExamOne requisition forms and shipped with biospecimens. Participants also completed the Patient-Reported Outcomes Measurement Information System-29 version 2 questionnaire (PROMIS-29 v2) through the Hugo Health interface within 24 hours of biospecimen collection.

### Biospecimen collection

Whole blood samples were collected in lithium-heparin-coated vacutainers (BD 367880, BD Biosciences) from participants’ homes or agreed upon locations, by ExamOne phlebotomists (Quest Diagnostics). Biospecimens were shipped overnight at regulated temperatures to Yale University in New Haven, CT on the day of collection. Upon receipt, collection tubes were de-identified according to protocol and study identifiers were provided. Samples were processed thereafter and within 48 hours of collection.

Biorender^54^ was used to create a graphical schematic on study design, cohorts and assays.

### Plasma and PBMC isolation

Plasma and peripheral blood mononuclear cells (PBMC) were isolated following protocols published previously^22^. Briefly, whole blood samples were centrifuged at 650g for 10 minutes at room temperature without brake. Plasma from each participant were pooled together to a single 15 mL polypropylene conical tube, aliquoted and stored at - 80℃. PBMCs were subsequently isolated using SepMate tubes (StemCell) following manufacturer’s instructions. Freshly isolated PBMCs were used for downstream flow cytometry analyses or were gradually cryopreserved using a freezing container and then transferred to liquid nitrogen tanks for long-term storage.

### Anti-SARS-CoV-2 nucleocapsid (N) antibody testing

The EUA-cleared Elecsys Anti-SARS-CoV-2 immunoassay (Roche Diagnostics, Indianapolis) was used to independently determine prior history of SARS-CoV-2 infection among participants at the Yale-New Haven hospital clinical research laboratory. Plasma aliquots were analyzed using this double-antigen sandwich (DAGS) immunoassay that measures total high affinity antibodies directed against the viral N protein. The COBAS e801 platform (Roche Diagnostics, Switzerland) was used according to the manufacturer’s instructions as validated earlier^25^.

Both self-reported infection history and the results of this assay were used to determine a participant’s history of SARS-CoV-2 infection. Participants were classified as infection positive if they reported a past infection but had no detectable anti-N antibodies, or if they had detectable anti-N antibodies regardless of not reporting a prior infection.

### Flow cytometry

Flow cytometry was performed on an Attune NxT Flow Cytometer (Thermo Fisher) using NxT v 5.31.0 software as described previously^22^. In brief, 1 to 2 million freshly isolated PBMCs were plated in round-bottom 96 well plates for surface and intracellular staining. Dead cells were stained using Live/Dead Fixable Aqua (ThermoFisher) on ice followed by a wash and Fc receptor blocking (Human TruStain FcX™, Biolegend). A total of 30 fluorophore conjugated antibodies were used in three different combinations for surface staining to define myeloid, B and T cell lineages. Quantitation of intracellular cytokine staining with stimulation was executed by incubating cells in 1× cell stimulation cocktail (eBioscience) without protein transport inhibitor for 1 hour in 10% FBS cRPMI followed by 4 hour incubation in 1x stimulation cocktail with protein transport inhibitor(eBioscience) in 10% FBS cRPMI at 37°C. Cells were permeabilized using 1× permeabilization buffer from the FOXP3/Transcription Factor Staining Buffer Set (eBioscience) at 4 °C and stained after Fc receptor blocking. FlowJo v.10.8 (BD) was used for data analysis. All the markers and gating strategies were as described previously and noted in the Flow Repository (ID: FR-FCM-Z8FZ) ^22^.

### SARS-CoV-2 antibody specific immunoassays

Enzyme-linked immunosorbent assays (ELISA) were run as described earlier ^55^. Briefly, 96-well MaxiSorp plates (Thermo Fisher Scientific, #442404) were coated at a concentration of 2 μg/ml in PBS with recombinant SARS-CoV-2 S protein (50 μl per well; ACROBiosystems, #SPN-C52H9-100 μg) or RBD (ACROBiosystems, #SPD-C52H3-100 μg) or nucleocapsid protein (NUN-C5227-100 μg, ACROBiosystems) and incubated overnight at 4°C. After removing the coating buffer, the plates were blocked with 200 μl of blocking solution (PBS with 0.1% Tween-20 and 3% milk powder) for 1 hour at room temperature (RT). Plasma samples were diluted 1:1500 (for Anti-Spike and Anti-RBD) and 1:400 (for Anti-N) in buffer (PBS with 0.1% Tween-20 and 1% milk powder), and 100 μl of the diluted plasma were added to the wells for 2 hours at RT. A standard curve was generated using serially diluted Human anti-Spike [SARS-CoV-2 Human Anti-Spike (AM006415) (Biolegend, #938602)] and anti-nucleocapsid SARS-CoV-2 human anti-nucleocapsid (1A6) (ThermoFisher Scientific, #MA5-35941). After three washes with PBS-T (PBS with 0.1% Tween-20), 50 μl of horseradish peroxidase-conjugated anti-Human IgG antibody (GenScript #A00166; 1:5000) diluted in buffer was added to each well and incubated for an hour. The plates were then developed with 100 μl of TMB (3,3’,5,5’-tetramethylbenzidine) Substrate Reagent Set (BD Biosciences #555214) and read at wavelengths of 450 and 570 nm.

### Linear peptide profiling & ELISA validation

Plasma aliquots were shipped to Serimmune on dry ice and the SERA assays were used to discover pathogen specific immunoglobulin G responses to linear peptides as described earlier ^22,56^. The published PIWAS method^57^ was used to identify candidate SARS-CoV-2 antigens and epitopes from the UniProt reference proteome (UP000464024). Briefly, the 12mer peptide sequences captured for each sample were broken down into 5- and 6-mers (k-mers) and tiled onto the proteome. An enrichment for each *k*-mer was then calculated as described ^57^ based on the frequency of that kmer in the dataset relative to the frequency expected by random chance. Outliers were identified by comparing the enrichment to a large pre-pandemic cohort (*n* = 1,500). An outlier threshold and outlier sum statistic were then derived, and a p value was calculated by comparing the observed outlier sum statistic to the null distribution. Outlier sum threshold was set to the 99.5th percentile value of all positions with FDR-adjusted *p* > 0.001. All sequence positions that exceeded both thresholds were identified, and adjacent positions were merged into regions representing candidate epitopes on the protein.

### ELISA Validation

96-well MaxiSorp plates (Thermo Scientific #442404) were coated with 200 ng per well of recombinant EBV gp42/ protein (MyBioSource #MBS430545) in PBS and incubated overnight at 4 °C. Plates were emptied and incubated with 3% Omniblock non-fat dry milk (American Bioanalytical #AB10109-00100) in PBS with 0.1% Tween 20 (Sigma-Aldrich, #P7949) for 2 hours at RT and then overnight at 4°C. Plates were washed 3× with 200 ul wash buffer (PBS 0.1% Tween 20). Samples and a serial dilution curve of monoclonal antibody against EBV gH/gL/gp42 (MyBioSource #MBS430548) were diluted in 1% Omniblock non-fat dry milk in PBS and added to the plate to incubate for 1 hour at RT. Plates were washed 3× with a wash buffer. Goat anti-human IgG Fc HRP (Sigma Aldrich, #AP112P) diluted 1:5000 or goat anti-mouse IgG Fc HRP (Southern Biotech #1030-05) diluted 1:6000 in 1% Omniblock non-fat dry milk in PBS was added to the plates and incubated for 50 minutes at RT. Plates were washed 5×. Plates were developed with 50 μl of TMB Substrate Reagent Set (BD Biosciences #555214) and the reaction was stopped after 5 min by the addition of 2 N sulfuric acid. Plates were then read at a wavelength of 450 nm.

### Immunoglobulin Isotyping & Autoantigen arrays

Plasma aliquots were shipped to Eve Technologies on dry ice. Two assays were run to determine the concentrations of the different Ig subtypes and IgG isotypes in the plasma, namely, Immunoglobulin Isotyping 6-Plex Custom Assay (IgG1, IgG2, IgG3, IgG4, lgA, and lgM) and the Human-IgE-1-Plex-Custom-Assay.

Multiplex IgM and IgG autoantibody reactivities were analyzed at the Microarray Core facility at the University of Texas Southwestern Medical Center, Dallas, TX, USA as described previously ^58,59^. Plasma aliquots were shipped to the facility on dry ice. A total of 120 human antigens, autoantibodies against which have been associated with various immune-related diseases or allergic disorders were included, namely, ACE2, Aggrecan, Albumin, α Fodrin, Amyloid β(1-40), Amyloid β(1-42), AQP4, BAFF, BCOADC-E2, BPI, Calprotectin/S100, CD4, CD40, CENP-A, CENP-B, collagen I, collagen II, collagen III, collagen IV, collagen V, Complement C1q, Complement C3, Complement C4, Complement C5, Complement C6, Complement C7, Complement C8, Complement C9, CRP, Cytochrome C, DFS70, dsDNA, EJ, Factor B, Factor H, Factor I, Factor P, fibrinogen Type I-S, fibronectin, GAD65, GBM, genomic DNA, Gliadin, gp210, GP2, H/K-ATPase, histone, histone H1, histone H2A, histone H2B, histone H3, HSPG, IA-2, IF, IFNγ, IL-6, IL-12/NKSF, IL-17A, Jo-1, KS, KU (P70/P80), La/SS-B, Laminin, LC1, LKM 1, LPS, lysozyme, M2, MBP, MDA5, Mi-2, mitochondrion, MPO, myosin, Nrp1, nucleolin, nucleosome, Nup62, NXP2, OGDC-E2, P0, P1, P2, PCNA, PDC-E2, PL-7, PL-12, PM/Scl-75, PM/Scl 100, PR3, proteoglycan, prothrombin, Ro/SS-A(52 kDa), Ro/SS-A(60 Kda), SAE1/SAE2, Scl-70, SLA/LP, Sm, Sm/RNP, SmD, SmD1, SmD2, SmD3, SP100, SRP54, ssDNA, Tau, thyroglobulin, TIF1γ, TLR4, TNFα, TPO, tTG, U1-snRNP 68/70kDa, U1-snRNP A, U1-snRNP C, U-snRNP B/B’, Vimentin, Vitronectin and ß2-Glycoprotein 1) were printed on 16 pad nitrocellulose Fast Analyte Scanning Technology (FAST) slides. Eight positive control proteins (human IgM and IgG, anti-human IgM and IgG) were also imprinted on the arrays as positive controls. Briefly, DNAse I (Thermo Fisher, #AM2222) treated plasma samples were applied onto the autoantigen arrays at 1:50 dilution. Binding was detected using cy5-labeled anti-human IgM and cy3-labeled anti-human IgG antibodies (Jackson ImmunoResearch Laboratories, #IgM 109-606-129, #IgG-109-166-098), and the array slides were scanned at wavelengths of 532 nm for and 635 nm for cy5 and cy3 respectively using a Genepix 4400A scanner. Genepix Pro v7.0 software (Molecular Devices) was used to analyze the image and generate the genepix report (GPR) files. The net signal intensity (NSI) values for each antigen were calculated by subtracting the negative control values. Normalized NSI values calculated using built-in positive controls were used for case-control analyses. Signal-to-noise ratios (SNR) for each antigen were calculated using the formula: (foreground median value – background median value)/standard deviation. Antigens with SNR<3 in more than10% of all samples were excluded. Log_2_ fold-change (FC) values between cases and controls were calculated.

### Multi-target soluble plasma factor analysis

Plasma aliquots stored at −80 °C were shipped to Eve Technologies on dry ice. All the samples were run in the same batch to avoid batch effects. The analytes covered by the following panels were assayed: Human Cytokine/chemokine 96-Plex Discovery Assay (HD96), Human Cytokine/chemokine Panel 4 12-Plex Discovery Assay(HDIV12), Human Adipokine 5-Plex Discovery Assay (HDADK5), Human Cardiovascular Disease Panels 2 and 3 (HDCVD8, HDCVD9), Human Complement Expanded Panel 1 9-Plex Discovery Assay (HDCMPEX1), Human MMP and TIMP 13-Plex Discovery Assay (HMMP/TIMP-S,P), Human Neuropeptide 5-Plex assay (HNPMAG-35K), Human Soluble Cytokine Receptor 14-Plex Discovery Assay (HDSCR14), Multi-Species TGF-β 3-Plex Discovery Assay (TGFβ1-3), Human Pituitary Panel 1 7-plex (HPTP1), Multispecies Hormone Panel 5-Plex (MSHMAG-21K) and Arginase-1.

To quantitate the total plasma testosterone levels, competitive ELISAs were performed as per manufacturer’s recommendations with the exception of the dissociation time which was increased to 30 mins to ensure complete release of protein bound testosterone (Thermo Fisher, #EIATES). Only analytes with less than 20% missing values were included in the analyses.

### SPEAR SARS-CoV-2 Spike Protein Immunoassay

The SARS-CoV-2 S1 antigen was measured in plasma samples using the SPEAR SARS-CoV-2 Spike Protein Immunoassay, developed and performed by Spear Bio, Inc. (Woburn, MA). The SPEAR immunoassay employs two specially designed DNA probes conjugated to target-specific antibodies that undergo a two-step successive polymerase extension reaction when bound to the target analyte, producing a unique DNA strand for analyte quantification with extremely low background. The assay uses antibodies specific to two distinct epitopes within the S1 subunit (GenScript, #A02052, #A02058). Plasma samples first underwent a reduction treatment to release S1 from potentially bound-blocking antibodies. In brief, plasma samples were incubated with SPEAR-specific reducing reagent for 15 minutes at 37°C, followed by the addition of a SPEAR-specific reduction quenching solution before incubation with the probes. The treated samples were then diluted tenfold in the assay diluent and incubated with S1-specific antibody-oligonucleotide probes for 2 hours at 37°C. This was followed by a 30-minute reaction to convert the presence of the analyte into DNA signal strands. The DNA signal strands were quantified through qPCR using the QuantStudio 12K Flex (ThermoFisher) with primers and qPCR probes specific to the DNA signal strand sequence.

Standard curves were generated from a stock solution S1 spike protein spanning from 0.0625 – 100,000 fM. Results from standard curves were used to generate sigmoidal four parameter logistic (4PL) fits in GraphPad Prism (v 10) software. Sample concentration results were calculated from run specific 4PL fit, multiplied by dilution factor. Lower limit of detection (LLoD) was calculated as the concentration corresponding to 2.5 standard deviation + mean zero in signal (18 replicates per run). Lower limit of quantification (LLoQ) was derived from the sample precision profile [measured concentration vs. concentration coefficient of variation (CV)] as the measured concentration at which a sample precision fit intersects at 20% concentration CV. The LLoQ and the LLoD values were calculated to be 20.5 fM and 5.64 fM as a mean of 6 runs. Values below LLoQ were replaced by the assay LLoD value and all values below LLoD were imputed as zero. The values were then transformed using the natural logarithm (ln(x + 1)) for comparisons. Analysis between groups was performed through a generalized regression model for zero inflation distribution (ZI Poisson) applying Tukey’s HSD to each pair.

To further validate the S1 assays, independent SPEAR immunoassays were performed to detect the full-length protein. This assay utilized oligonucleotide probes conjugated to antibodies targeting both the S1 (GenScript, #A02052) and S2 (R&D Systems, # MAB11362-100) subdomains. The S1 subunit binding antibody was common to both the assays.

### General Statistical Analyses

The association of variables between case and control cohorts and among the four groups were assessed using Fisher’s exact test for categorical variables. For pair-wise comparisons, Mann-Whitney tests with Benjamini-Hochberg correction for multiple testing were used. For four-group comparisons, Kruskal-Wallis tests followed by Conover’s post-hoc test with Benjamini-Hochberg method for multiple comparisons correction were applied to continuous variables. Post hoc tests were also performed using Dunn’s test with Bonferroni–Holm’s method to adjust for multiple comparisons. Both one and two-sided p values ≤ 0.05 were considered statistically significant. Spearman’s rank correlation tests were utilized to analyze the relationships between cell populations, antibody levels, anti-SARS-CoV-2 antibody levels, the number of vaccinations, days post-vaccination, days post-exposure and in assay validations.

### Linear modeling

Generalized linear modeling was performed linear regression with outputs of model coefficients for each variable and associated p-values ≤ 0.05 were considered statistically significant. All analysis was performed using R. Standard generalized linear model function listed is below.

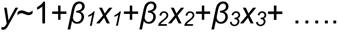

### Machine Learning Analysis

To predict PVS and control outcomes, we utilized a two-step approach involving weighted gene co-expression network analysis (WGCNA) and the construction of a generalized LASSO linear model^31,60^. Initially, we preprocessed the data via quantile normalization to address outliers and skewness, with average ranks assigned in case of ties. Due to the presence of highly correlated features, we applied WGCNA to identify clusters of features with similar expression patterns. Specifically, we set the soft thresholding power parameter to 7, which is crucial for transforming the raw correlations into adjacency values, by maintaining the highest mean connectivity and achieving a model fit higher than 0.8. Using this selected parameter, we computed the network adjacency based on pearson correlation within a signed network. We then constructed a topological overlap matrix (TOM) and converted it into its corresponding dissimilarity measure. Hierarchical clustering was performed on this measure with ward.D2 linkage, and the dendrogram was pruned to enforce a minimum cluster size of 5. This process allowed us to identify 6 distinct modules.

We built the LASSO model using raw features from module 1, a large module with numerous features that was not further clustered, along with the eigengenes (first principal component) of the other 5 modules. To compensate for small sample size and assess model performance, we conducted nested cross-validation with both inner and outer loops set to 10, and adjusted the classification threshold to 0.65 to combat class imbalance. Additionally, a permutation test was performed by shuffling the PVS label within the infection and non-infection group, demonstrating the fitted model performed better than random chance. We then refitted the LASSO model using all available data and applied the min criterion rule to select important features. Given that LASSO does not inherently provide p-values due to its regularization nature, we also assessed the marginal significance of each individual feature using logistic regression. To better understand model performance, t-SNE, or t-distributed Stochastic Neighbor Embedding, was used to create low-dimension plots of the LASSO variable space.

Lastly, to ensure model robustness, a secondary workflow was utilized to examine variable importance. For this, a random forest model was created utilizing nested cross-validation and SMOTE up-sampling to compensate for class imbalance. Random forest permutation importance was then used as a metric to examine the top features.

