## Supplementary Figures for "Immunological and Antigenic Signatures Associated with Chronic Illnesses after COVID-19 Vaccination"

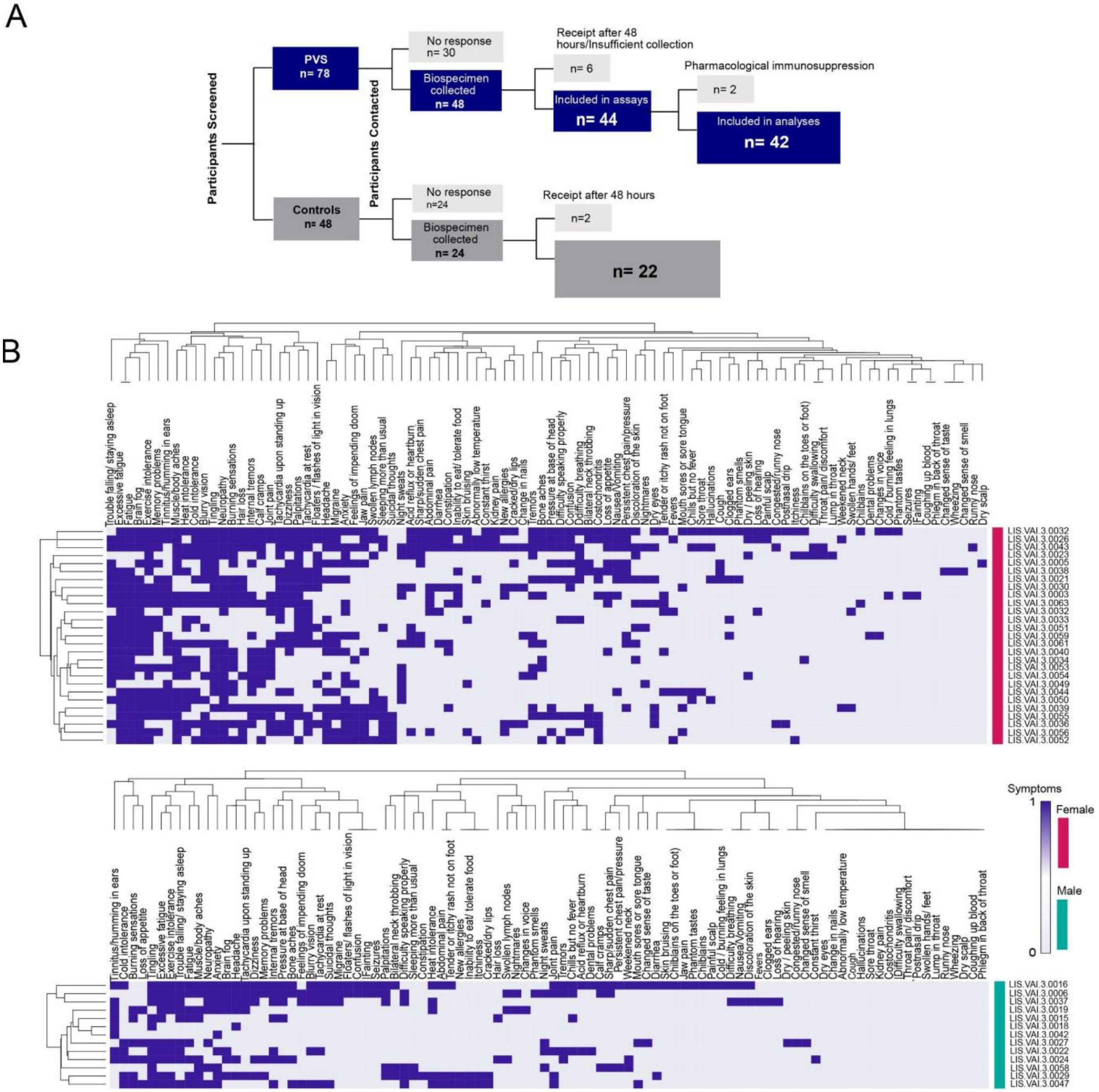


**Figure S1: Hierarchical clustering of PVS symptoms. A.** A flow diagram summarizing the steps leading to the final set of participants included in the assays and analysis, from initial screening to the final selection. **B.** Dendrograms based on hierarchical clustering based on self-reported symptoms at recruitment in female and male participants. Symptom percentages are shown in Table S1.

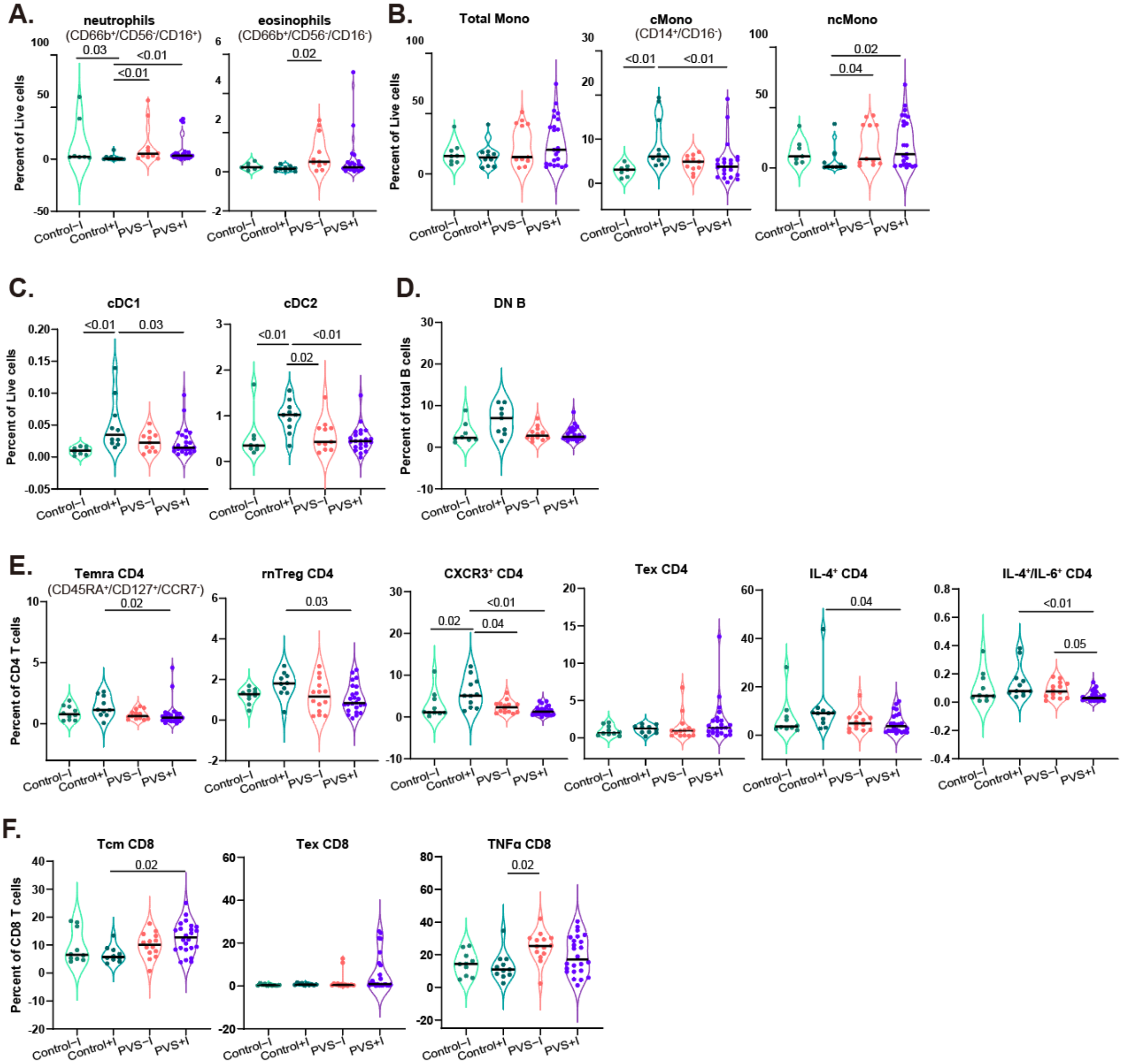


**Figure S2: Immune cell feature of myeloid and lymphoid cells in PVS patients grouped by SARS-CoV2 infection history. A-C.** Violin plots of myeloid peripheral blood mononuclear populations (PBMCs) plotted by the group as percentages of respective parent populations (live cells). **D.** Violin plots of double negative B cells from PBMCs plotted as percentages of total B cells. **E.** Violin plots of various CD4 T cell subsets and cytokine-producing CD4 T cell subsets. **F.** Violin plots of central memory CD8 T cells and TNFα-producing CD8 T cells.

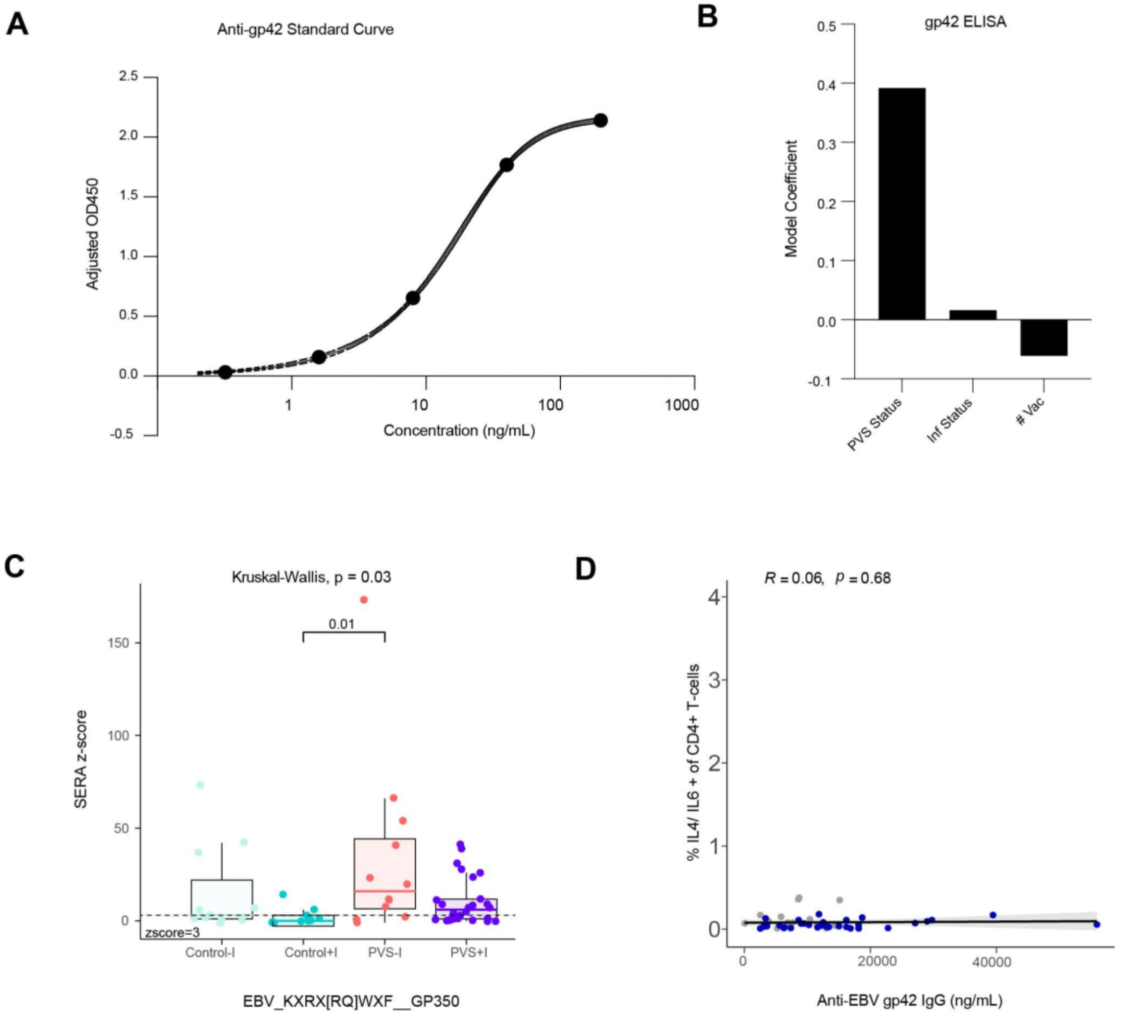


**Figure S3: Responses against herpesvirus and EBV A.** Mean absorbance (OD450nm) plotted against serial dilutions of monoclonal IgG antibody against EBV gp42 to generate a standard curve. Best fit was determined using asymmetrical sigmoidal five-parameter least-squares fit. Projected antibody concentrations were interpolated using this fit. Showing one of four replicates. R-squared = 1, Sum of squares = 4.07e-005. **B.** Coefficients from linear models are reported for anti-gp42 antibody responses. Model predictors are reported along the x-axis and included age, sex (categorical), PVS status (categorical), infection status group (categorical), and number of vaccinations at blood draw (# Vac). Significant predictors ($p \leq 0.05$) are plotted in blue. Detailed model results are reported in Table S3. **C.** SERA-derived *z* scores for the gp350 motif KXRX[RQ]WXF among EBV-seropositive individuals only, plotted by group, $n = 11$ (Control - I), $n = 9$ (Control + I), $n = 12$ (PVS - I), $n = 26$ (PVS + I). The dashed line represents the *z*-score threshold for epitope positivity defined by SERA. **D.** The relationship between plasma concentration of IgG against EBV gp42 and the percentage of IL4/IL6 + CD4+ T cells (of total CD4+ T cells) for participants. Correlation was assessed using Spearman's correlation. The black line shows linear regression, and the shading shows the 95% CIs.

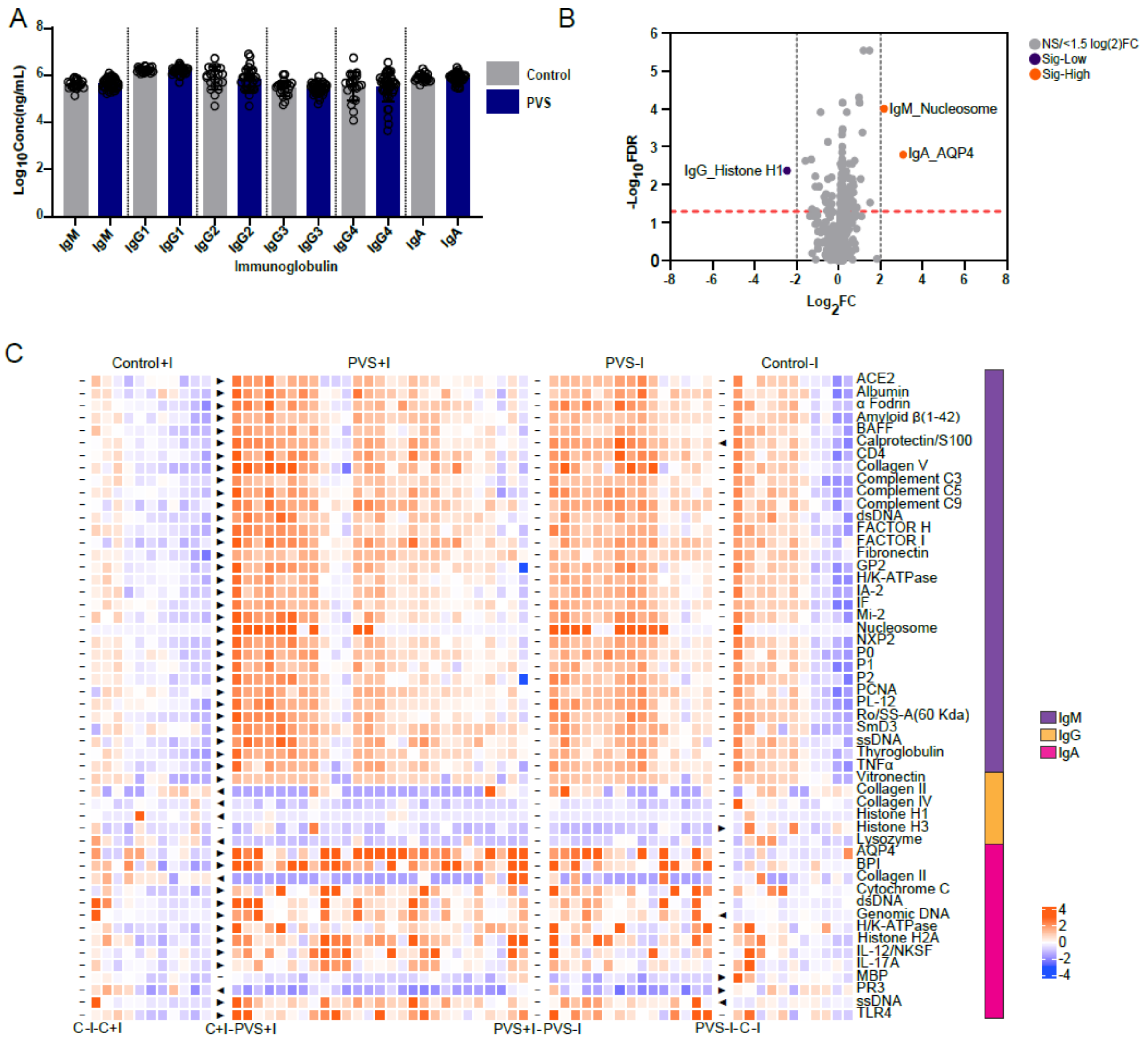


**Figure S4: Autoantibodies in circulation across cohorts. A.** Bar graphs of antibody subtype and isotype titers in circulation are plotted based groupwise. **B.** A volcano plot showing differences in autoantibody titers between PVS and controls. The orange horizontal dotted line indicates the $\log_{10}$ FDR (Benjamini Hochberg method) of 0.05 which is 1.3. The gray vertical dotted lines indicate a $\log_2$fold change of 2 which is equal to a fold change of 4. All autoantibodies not differing significantly are highlighted in gray. Autoantibodies with significantly different titers are shown in violet (lower in PVS) or orange (higher in PVS). Kruskal-Wallis tests with Conover post-hoc tests for multiple comparisons correction were used. Adjusted p-values < 0.05 are indicated. Detailed analyses results are shown in Tables S4 and S5.

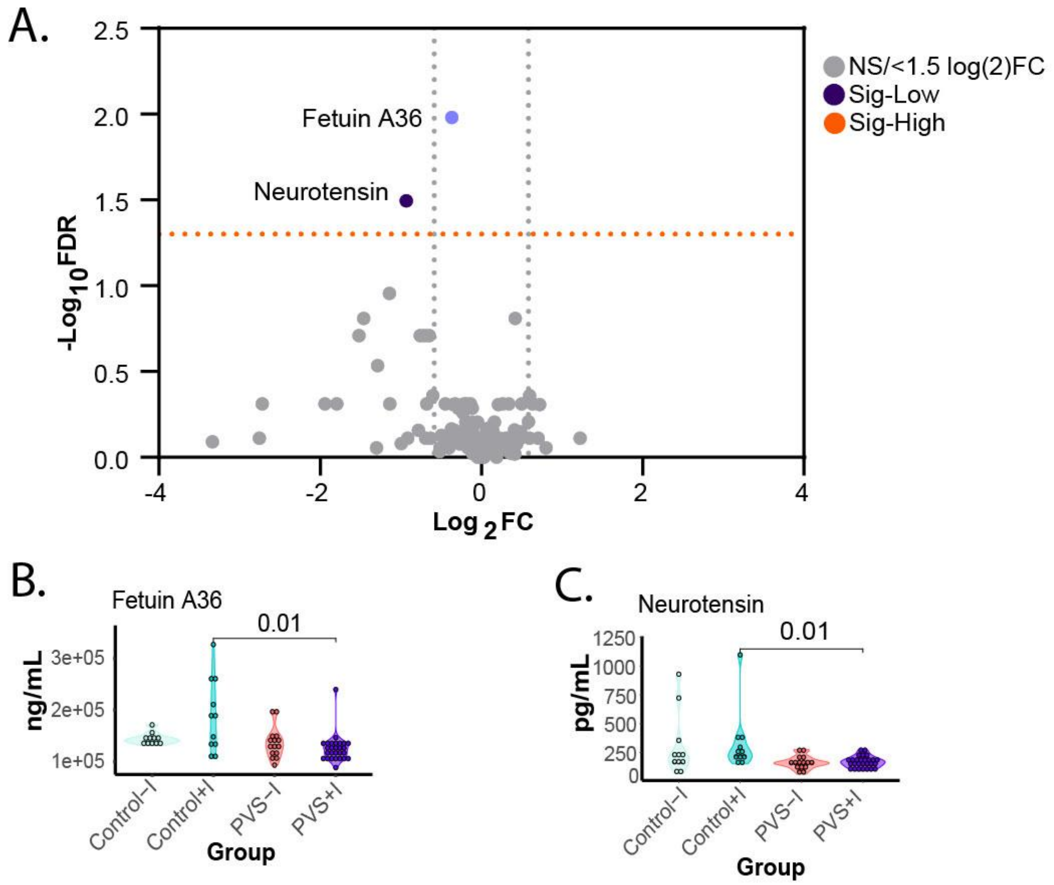


**Figure S5: Soluble plasma factors across cohorts. A.** A volcano plot showing differences in soluble factors between PVS and controls. The orange horizontal dotted line indicates the $\log_{10}$ FDR (Benjamini Hochberg method) of 0.05 which is 1.3. The gray vertical dotted lines indicate a $\log_2$fold change of 0.585 which is equivalent to a fold change of 1.5. All soluble moderators showing no differences are highlighted in gray. Soluble modulators with significant differences in concentration are either indicated in violet for fold change lower than 1.5 and purple for fold change greater than 1.5. **B-D.** Violin plots of circulating factors that differ significantly among the control-I, control+I, PVS-I and PVS+I subgroups. Kruskal-Wallis tests with Conover post-hoc tests for multiple comparisons correction were used. Adjusted p-values < 0.05 are indicated. Detailed analyses results are shown in Table S6 and S7.

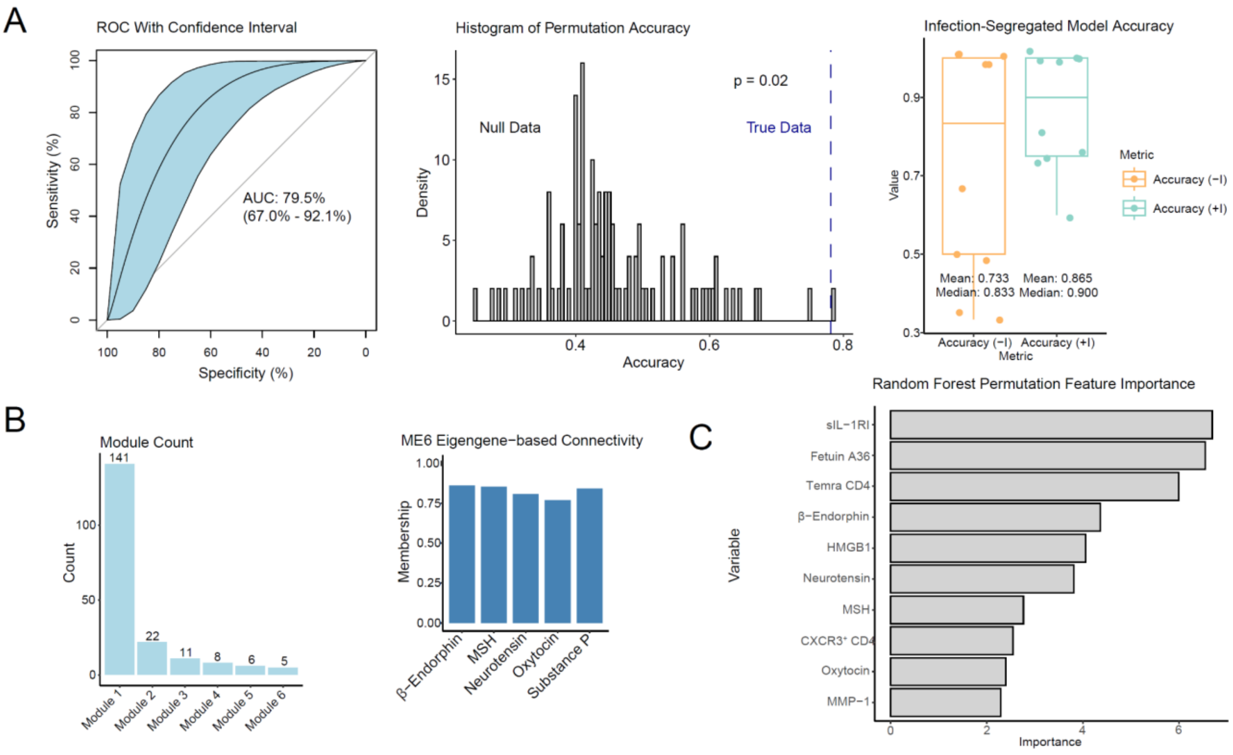


**Figure S6: Additional Machine Learning Results. A.** LASSO ROC curve, binomial smoothing applied, with 95% confidence interval ROC. Relevant AUC and 95% confidence interval. LASSO permutation test with 100 iterations, with dashed blue line denoting the true accuracy, and resultant p-value. Infection-segregated per-fold model accuracy, created by splitting the test data per-fold by infection status. For each, mean and median fold accuracy is provided. **B.** Histogram showing the size of all WGCNA modules. Module 6 eigengene-based connectivity. **C.** Permutation importance scores for the top 10 features selected by a random forest model.
